# Genomic-Relatedness Matching Expands Population Coverage, Improves Power, and Reduces Bias in Genetic Association Analyses

**DOI:** 10.64898/2026.05.14.26353140

**Authors:** Dhruva Jaishankar, Tamara Gjorgjieva, Jonathan Jala, Jeffrey Swigert, Alexander Strudwick Young, Daniel J. Benjamin, David A. Cesarini, Patrick Turley

**Affiliations:** Anderson School of Management, University of California Los Angeles, Los Angeles, CA, USA; Department of Genetics, Stanford University, Stanford, CA, USA; National Bureau of Economic Research, Cambridge, MA, USA; Center for Economic and Social Research, University of Southern California, Los Angeles, CA, USA; Human Genetics Department, UCLA David Geffen School of Medicine, Los Angeles, CA, USA; Department of Economics, New York University, New York, NY, USA; Department of Economics, University of Southern California, Los Angeles, CA, USA

**Author notes:** Correspondence to Patrick Turley.

## Abstract

We introduce a novel approach, Genomic-Relatedness-Matched Association (GRMA) studies, as an alternative to genome-wide association studies (GWAS). GWAS are typically restricted to samples of mostly unrelated individuals with a single, shared continental ancestry and nevertheless can still be biased by gene-environment correlation and assortative mating. In contrast, GRMA can be implemented in ancestrally diverse samples—retaining individuals of mixed or underrepresented ancestries and eliminating the need to assign labels to ancestry groups—and can reduce bias relative to standard GWAS. GRMA matches each individual to a group of controls whose pairwise relatedness with the individual exceeds a user-specified threshold. It generates SNP-level summary statistics based on within-group associations. In applications using the UK Biobank and All of Us data, we find that GRMA compares favorably to GWAS methods in terms of bias, precision, and population coverage. GRMA enables several novel findings; for example, we find that “genetic nurture” is unlikely to be an important source of genome-wide bias in population GWAS of body mass index, height, and educational attainment. The method is computationally efficient and supported by open-source software, facilitating its application in large-scale scientific and health-related studies.

## INTRODUCTION

Genome-wide association studies (GWAS) of complex traits in humans have typically relied on samples of unrelated individuals drawn from a single continental ancestry, typically European^1,2^. While such population GWAS have delivered many discoveries^1^, they can produce substantially biased estimates.^3–5^ Standard GWAS protocols incorporate safeguards intended to reduce confounding, but these approaches do not always succeed at eliminating all biases from interpersonal genetic effects ^6,7^ (often called “indirect effects”), population stratification^4,8,9^, and assortative mating^10^. For some phenotypes in some populations, the residual bias can be substantial^11^.

Family-based GWAS approaches, such as sibling or trio GWAS, are an effective strategy for addressing these biases. Currently, however, family-based GWAS approaches have very limited power, both because the sample sizes of genotyped families are relatively small and because, holding sample size fixed, within-family GWAS is less statistically powerful than population GWAS^3,4,11,12^.

Here, we introduce the **Genomic-Relatedness-Matched Association (GRMA) study**. GRMA has two key attractive features. First, GRMA can reduce bias relative to population GWAS—but can do so while retaining more power than family-based GWAS. Second, GRMA can be implemented without restricting the estimation sample to individuals with a single, shared genetic ancestry. Indeed, GRMA does not require the assignment of *any* ancestry labels. This has two advantages: it increases statistical efficiency by retaining individuals of mixed or underrepresented ancestries in the analyses, and it sidesteps issues raised by the assignment and use of ancestry labels, most notably the concern that they are mistakenly equated with socially constructed racial classifications^13–16^.

GRMA is based on a logic familiar from the literature on matching methods for causal inference: for each individual, a *relatedness group* is generated consisting of controls whose genetic relatedness with the individual meets some relatedness criterion. Operationally, phenotypes and genotypes are residualized within relatedness groups, and SNP-level estimates are obtained by ordinary least-squares regression. Intuitively, GRMA can reduce bias because members of a relatedness group draw their alleles from more similar groups than unrelated individuals in a population GWAS. Including diverse ancestries in the analysis does not increase bias substantially because genetic associations are identified entirely from variation among individuals within a relatedness group, individuals who generally share similar genetic ancestries.

In this paper, after developing the theoretical underpinnings of GRMA and describing its computational implementation, we apply it to three phenotypes available in the UK Biobank^17^ (UKB): body mass index (BMI), height, and educational attainment. Using GRMA with third-degree relatives, with no detectable increase in bias relative to a sibling GWAS, we obtain power equivalent to a sibling GWAS with 145,646 siblings for BMI, 158,957 siblings for height, and 237,100 siblings for educational attainment. In contrast, after combining data from many cohorts (the UKB alone contains only approximately 41,000 siblings), the largest published sibling GWAS to date^12^ reported smaller sibling sample sizes (140,883 for BMI, 149,174 for height, and 128,777 for educational attainment) than we obtained from UKB alone.

GRMA’s additional power generates new insights. For example, prior family-based work has failed to reject the null that the genetic correlation between BMI and educational attainment is zero; using GRMA-based polygenic indexes, we find a within-sibship association of moderate magnitude that is statistically distinguishable from zero. As another example, analysis of GRMA’s bias at different relatedness thresholds provides insights into the sources of bias in population GWAS: our results imply that “genetic nurture” is unlikely to be a major contributor to bias in the UKB for the phenotypes we study. We anticipate that GRMA will increase the value of large samples with genetic data and accelerate scientific discoveries that require precise estimates of causal genetic effects.

## RESULTS

### GRMA Framework

Consider a dataset with individuals indexed by *i* = 1,2, . . ., *N* and SNPs indexed by *k* = 1,2, . . ., *K*. Let *x*_*ik*_ denote *i*’s genotype, *f*_*ik*_ denote the midpoint of *i*’s parents’ genotypes, and *y*_*i*_ denote *i*’s phenotype. The parameter of interest is the coefficient *β*_*k*_ that would be obtained from the trio-based regression

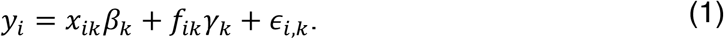

This coefficient is robust to assortative mating and all biases from gene-environment correlation, including population stratification and interpersonal genetic effects, because under Mendelian transmission, variation in genotype conditional on the parental midpoint is uncorrelated with the residual, *ϵ*_*i,k*_.^18–20^

In practice, the main barrier to estimating Equation (1) directly is that *f*_*ik*_ is not observed or reliably imputed in a sufficiently large sample. Biases are introduced when *f*_*ik*_ is omitted from the regression. Population GWAS mitigates the bias by including control variables, such as principal components, but these often do not fully eliminate the bias^11,20–22^.

GRMA instead identifies for each person a group of relatives, which we call the person’s *relatedness group*, and produces within-group estimates for each SNP. By estimating within relatedness groups, GRMA differences out all factors that are shared by individuals in the same group. When most of the confounding factors in a population GWAS are shared within groups, differencing out these factors will produce substantially less biased estimates than a population GWAS.

#### The Within-Relatedness-Group Model

We denote deviations from a variable’s relatedness-group mean using tildes. Demeaning both sides of Equation (1) at the relatedness-group level, we can write 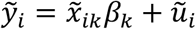, where *ũ*_*i*_ is a composite error 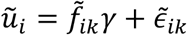. For each SNP *k*, the GRMA estimate is simply the ordinary least squares (OLS) coefficient from the regression of 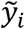 on 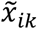.

The decomposition of *ũ*_*i*_ highlights two distinct sources of potential bias in GRMA estimates: (1) 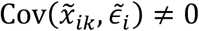, which arises when *ϵ*_*ik*_ includes the genotypes of relatives and therefore *i*’s outcome is affected by those genotypes (e.g., interpersonal genetic effects from siblings); and (2) 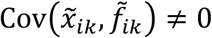, which arises when residual population confounds remain even within a relatedness group (e.g., admixture within extended families).

An important special case is when relatedness groups consist exclusively of full siblings. If *i* and *j* are siblings, then *f*_*ik*_ = *f*_*jk*_, so the 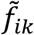 term vanishes in the demeaned model, leaving genetic effects from siblings as the only potential source of bias^18–20,23^. Since research to date has failed to produce credible evidence of sibling genetic effects of meaningful magnitude^20^, it is plausible that a GRMA analysis based only on siblings yields estimates with minimal bias: 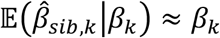.

#### Relatedness-Group Formation

Here, we outline the two-step algorithm used by GRMA to generate relatedness groups (for a more detailed description, see **Online Methods**).

First, genome-wide data are used to infer all pairwise genetic relationships between participants in the available sample. Each pair is assigned one of the following *relationship types*:

- Siblings (S),
- Parent-Offspring (PO),
- 2^nd^-degree relatives (2),
- 3^rd^-degree relatives (3),
- Unmatched (approximately unrelated) (U).

GRMA does not use relative pairs more distant than 3^rd^-degree because it is not always possible to reliably distinguish between 4^th^-degree and more distant relatives with genotype data alone.

Second, a relatedness group 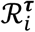 is formed for each individual *i* = 1,2 …, *N* and threshold *τ* = *S, PO*, 2,3 (and we omit the *τ* superscript whenever doing so does not cause ambiguity). The relatedness groups are formed iteratively. First, anyone with a sibling gets matched with all their siblings in the sample. Next, anyone unmatched who has a parent or offspring gets matched with all their parents and offspring. This is iterated for 2^nd^- and then again for 3^rd^-degree relatives (assuming the GRMA analysis is conducted at the 3^rd^-degree threshold). In this way, each person’s relatedness group consists of the reference individual and a group of people who are equally related to the reference individual. Also, this approach means that being in a relatedness group is reflexive 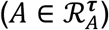, but not symmetric (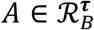 does not imply 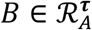) or transitive (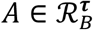 and 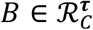 does not imply 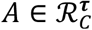). **Figure 1** illustrates relatedness-group formation using a simple example.

**Fig. 1.**
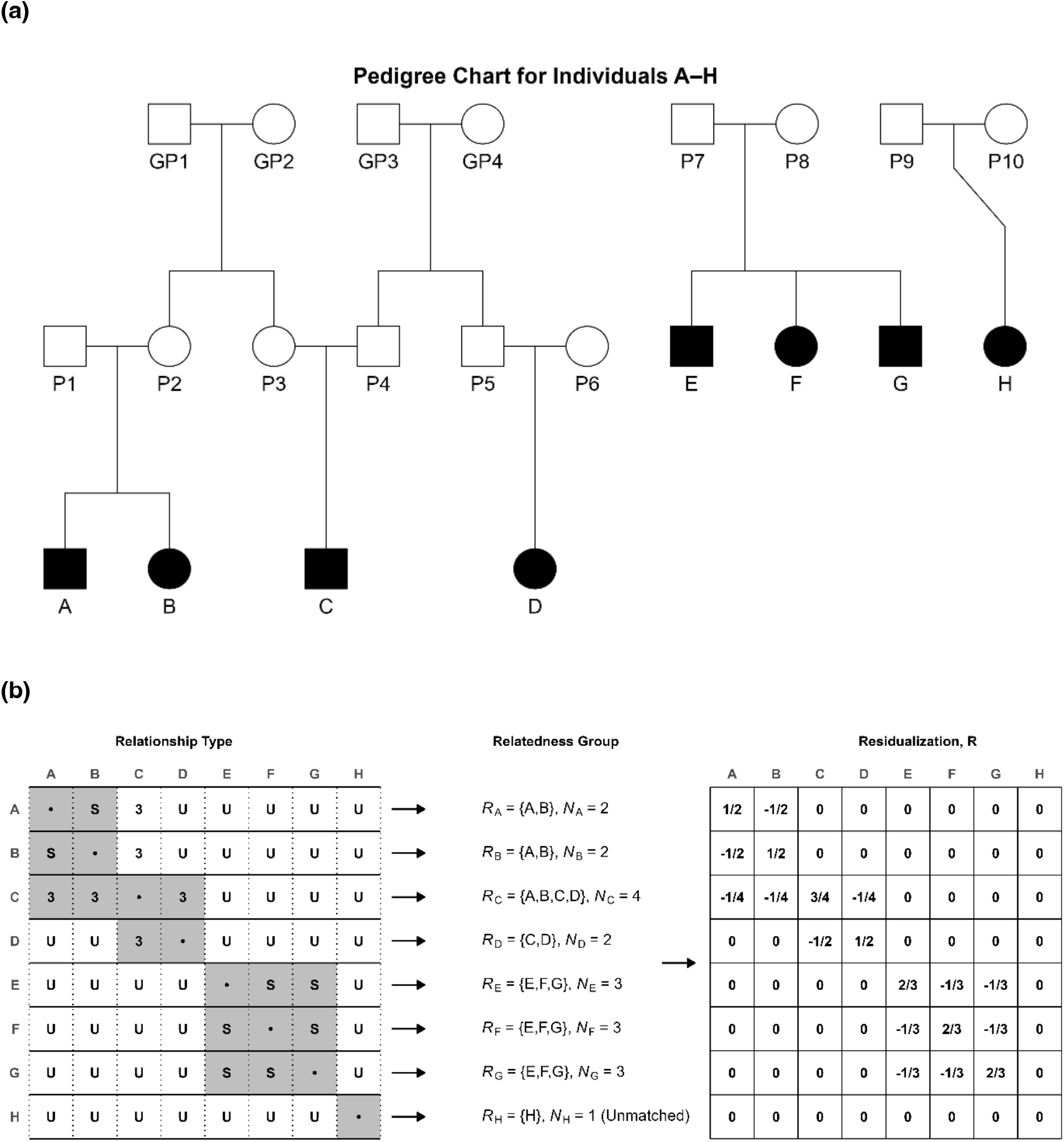
Illustration of Hierarchical Construction of GRMA Relatedness Groups and Residualization Matrix, R. **(a)** Schematic pedigree constructed to illustrate the GRMA grouping logic. **(b)** Illustration of how GRMA forms relatedness groups and the residualization matrix, **R**. The left matrix maps each pair of individuals to the degree of relatedness based on the pedigree in panel (a). (**⋅** : Self; **S**: sibling; **3**: 3rd-degree relative, **U**: unmatched.) Highlighted entries in a row show all individuals in the corresponding person’s relatedness group based on a 3rd-degree relative matching criterion. The middle panels list each relatedness group and group size explicitly. The right matrix shows the corresponding **R** matrix, which residualizes the phenotypic and genotypic data before the regression step. (See **Online Methods**.)

### GRMA Estimator and Inference

Omitting SNP subscripts and using capital letters in boldface to denote the length-*N* vectors, Equation (1) can be expressed compactly as ***Y*** = ***X****β* + ***F****γ* + ***ϵ***. GRMA is implemented by pre-multiplying ***Y*** and ***X*** by a residualization matrix ***R*** that demeans the data at relatedness-group level (see **Online Methods** and **Figure 1b**).

The GRMA estimator is OLS using the residualized variables:

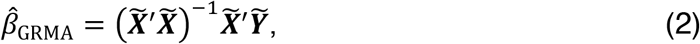

where 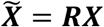 and 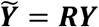.

Due to correlated residuals within and across relatedness groups (even after demeaning), the conventional formula for OLS standard errors is invalid. As shown in **Online Methods** and **Supplementary Note**, the sampling variance of 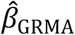 is:

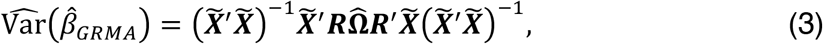

where 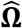 is an estimate of the variance-covariance matrix of ***Y***. In simulations, we find that the GRMA standard errors yield an accurate Type I error rate for null SNPs across a range of significance thresholds (see **Supplementary Figure 1** and **Online Methods**).

We provide an open-source Python command line tool that implements this procedure efficiently at biobank scale; the software is publicly available together with documentation (URLs in **Data and Code Availability**).

### Bias–Variance Tradeoff

In most applications of GRMA, researchers face a bias-variance tradeoff. On the one hand, a more permissive relatedness threshold improves precision by increasing the number of individuals included and by increasing the genotypic variance within each relatedness group. On the other hand, greater precision could come at the cost of greater bias.

The tradeoff can be formalized through a *modified mean-squared error* (MMSE) criterion, defined as:

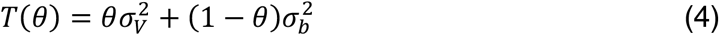

where 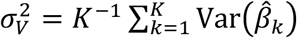 is the *genome-wide-mean sampling variance*, 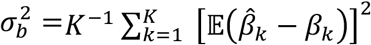 is the *genome-wide-mean squared bias*, and *θ* ∈ [0,1] is a weight parameter that describes a researcher’s aversion to sampling variance relative to bias. For *θ* = 0.5, *T*(*θ*) is the standard mean-squared error (MSE) criterion.

A natural way to estimate 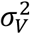 is to replace each summand 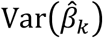 by its sample analog, 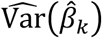, and calculate 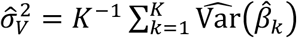. To estimate 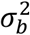, instead of using *β*_*k*_ as the target estimand, we use the expectation of the sibling GRMA estimator. We derive an estimator, 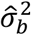, that depends only on the sibling-GRMA summary statistics and those obtained at the alternate relatedness threshold. Our estimators for 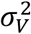 and 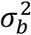 and the block-jackknife procedure we use to estimate standard errors are described in the **Supplementary Note** and **Online Methods**. Using these estimates, we can calculate ranges of *θ* for which a researcher may prefer one relatedness threshold over another.

### Application and Evaluation of GRMA

In UK Biobank^24,25^ (UKB) data, we apply GRMA to three phenotypes: body mass index (BMI), height in centimeters, and educational attainment in years of completed schooling. (See **Supplementary Tables 1-2**.) We apply standard quality-control genotype filters, resulting in a set of 828,208 SNPs. (See **Online Methods** and **Supplementary Table 3**.) We report two complementary comparisons. The first compares GRMA analyses conducted at all four relatedness thresholds: siblings (GRMA-S), parent–offspring (GRMA-PO), second-degree (GRMA-2^nd^), and third-degree (GRMA-3^rd^). For each phenotype, we characterize how the relatedness threshold impacts statistical power, polygenic prediction accuracy, and genome-wide bias. Next, we compare GRMA to a conventional population-based GWAS implemented in a non-overlapping sample of UKB subjects.

### Within-GRMA Comparisons

We begin by comparing summary statistics from GRMA analyses performed at different relatedness thresholds. To maximize comparability, we restrict all GRMA analyses to a common set of SNPs (**Online Methods**). To convey the main patterns in the data, our discussion below highlights the comparison between the GRMA-S and GRMA-3^rd^ estimates; complete results for all four relatedness thresholds are in **Supplementary Tables 4-8**.

Our estimation sample consists of all genotyped UKB participants (with no genetic-ancestry restrictions) who meet a set of standard quality-control filters (see **Online Methods**). We then use the population-structure-robust KING estimator^26^ to assign a relationship type to all pairs of individuals in the sample.

**Table 1** reports, for each phenotype and relatedness threshold, the number of individuals matched with at least one relative; individuals with no such relatives are defined as *unmatched*. Roughly 70% of individuals fall into this unmatched category, indicating that most UKB participants lack relatives at or above the 3^rd^-degree threshold. However, we find many more matched individuals at the 3^rd^-degree threshold compared to at the sibling threshold, illustrating GRMA’s value when stricter thresholds are underpowered.

**Table 1:** Number of Individuals Matched by Relatedness Threshold.

| Relatedness Threshold | # Individuals with Non-Missing Phenotype |  |  |
| --- | --- | --- | --- |
|  | BMI | Height | EA |
| **Siblings** | 40,960 | 41,019 | 40,331 |
| **Parent-Offspring** | 51,071 | 51,158 | 50,224 |
| **2<sup>nd</sup>-degree** | 68,478 | 68,619 | 67,345 |
| **3<sup>rd</sup>-degree** | 142,717 | 142,965 | 140,434 |
| **Unmatched** | 339,063 | 339,340 | 336,496 |
| **WB Unmatched** | 278,474 | 278,676 | 277,418 |

**Note.** Number of UK Biobank participants with at least one qualifying relative at each GRMA relatedness threshold or closer. “Unmatched” refers to all individuals who have **no** relatives at or above the 3<sup>rd</sup>-degree threshold; this group includes participants of all genetic ancestries. In contrast, the “WB Unmatched” sample is restricted to the subset of unmatched individuals who are classified by UKB<sup>26</sup> as “White British.” The label is assigned to those who self-report as White British and are not genetic ancestry outliers based on the first two principal components of the genotype data. The WB Unmatched sample has two useful properties. First, it is constructed using procedures that closely align with standard population-GWAS protocols for obtaining a genetically homogeneous estimation sample. Second, by construction, no individual in the WB Unmatched sample has a third-degree or closer genetic relationship with any individual included in a GRMA analysis, regardless of threshold.

The bottom row (WB Unmatched) reports the subset of unmatched individuals who are classified by the UKB as “White British”^17^, which we use as a prediction sample and for a population GWAS in later analyses. Of the roughly 140,000 individuals who have at least one 3^rd^-degree (or closer) relative, around 14,750 of them are not classified as being “White British,” meaning that these are individuals who would be omitted from a population GWAS due to their genetic ancestry^27^ but who can be included in a GRMA study since GRMA doesn’t use ancestry classifications. In other more diverse datasets, GRMA’s value can be even larger. For example, the All of Us sample has 57,045 individuals with at least one 3^rd^-degree or closer relative (less than half the number of such individuals in the UKB), and yet 35,755 of those individuals would be omitted from a European-genetic-ancestry population GWAS; GRMA includes all these individuals.

#### Statistical Power

For each phenotype, we evaluate the GRMA summary statistics in terms of statistical power using two metrics: the mean *χ*^2^ statistic and the mean sampling variance.

For all three phenotypes, the mean *χ*^2^ statistic increases substantially as the relatedness threshold becomes more permissive. To contextualize these increases, we compute the number of siblings that would need to be added to each GRMA-S analysis to inflate its mean *χ*^2^ statistic to that of the GRMA-3^rd^ results. Using the data in **Supplementary Table 7**, our projections indicate that the number of siblings would need to increase from 40,960 to 145,646 for BMI (356%), from 40,331 to 237,100 for educational attainment (588%), and from 41,019 to 158,957 for height (388%) (see **Online Methods**).

We also compare sampling variances across GRMA thresholds. **Figure 2** compares mean sampling variances in the GRMA-S and GRMA-3^rd^ analyses (for results at all four GRMA thresholds, see **Supplementary Table 8**). The ratio of GRMA-S to GRMA-3^rd^ mean sampling variances is similar across phenotypes, ranging from 3.4 (height) to 4.2 (BMI). We also find that GRMA-3^rd^ has substantially smaller sampling variance than an alternative family-based methods, snipar-Robust and snipar-Unified^23^, which use imputed parental genotypes from available first-degree relatives. (See **Online Methods** and **Supplementary Table 9**.)

**Fig. 2.**
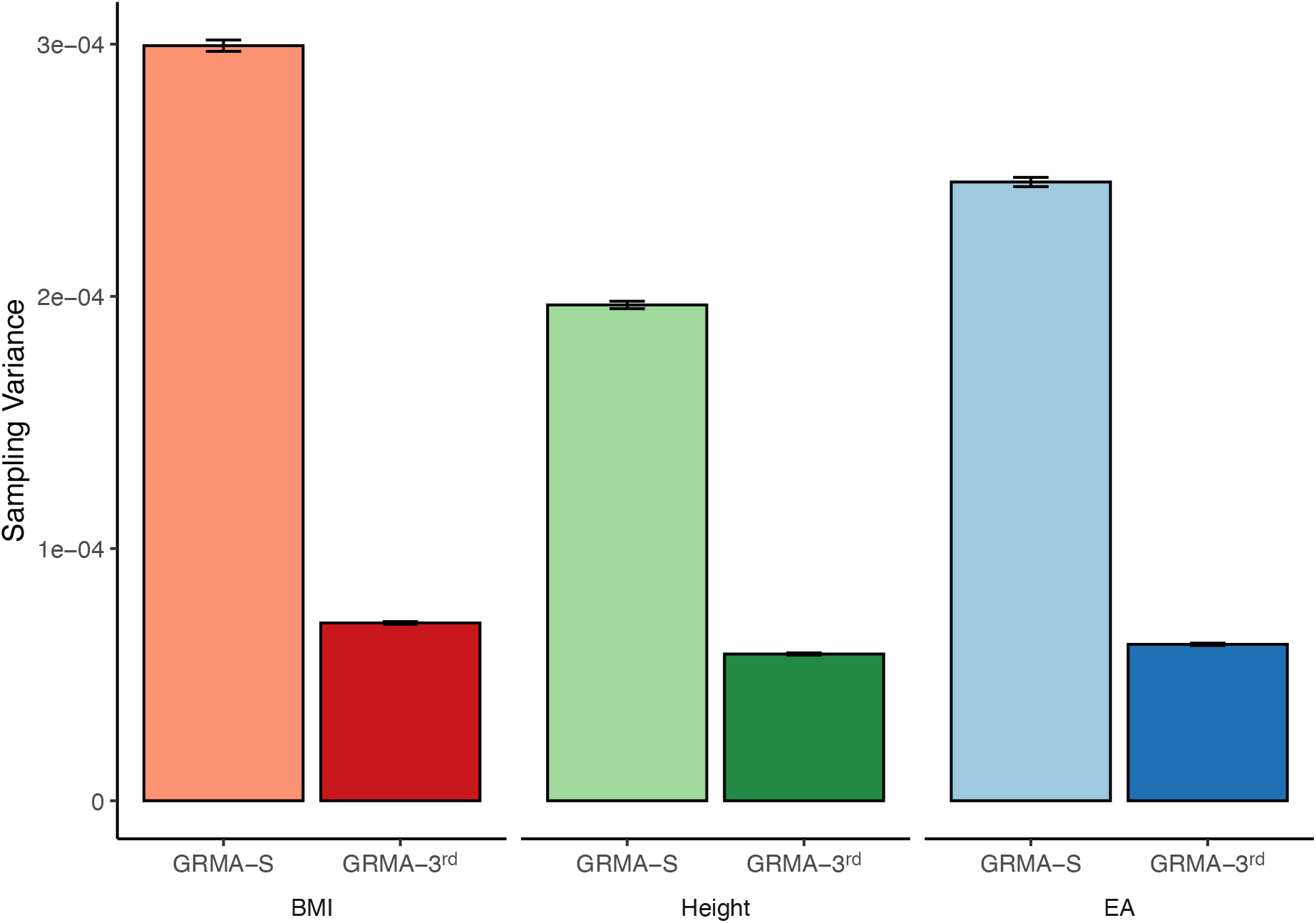
Mean Sampling Variance. Estimated sampling variance of GRMA-S and GRMA-3^rd^ for BMI, Height, and Educational Attainment (EA). Estimated as the mean squared standard error of the GRMA study summary statistics for a common set of approximately 830,000 HapMap3 SNPs. (See **Online Methods**.)

These comparisons show that GRMA-3^rd^ delivers far more precise estimates; the next question is whether that extra precision comes at the cost of materially greater bias.

#### Polygenic Prediction

While more precise estimates should increase the predictive power of polygenic indexes based on those estimates, this may be offset by increased bias. For each phenotype and each relatedness threshold, using GRMA summary statistics and the SBayesR^28^ method (**Online Methods**), we construct a set of polygenic indexes and compare their prediction accuracy. We do this in two non-overlapping holdout samples: (i) approximately 275,000 WB Unmatched individuals from the UKB (see **Table 1**), and (ii) approximately 12,800 siblings (with no genetic ancestry restrictions) in the All of Us Research Program^29^. By construction, the UKB sample does not overlap and is not closely related with any GRMA estimation samples. The siblings in the All of Us sample permit within-family analyses that are not vulnerable to sources of confounding that may be present in the UKB holdout sample.

For all three phenotypes, the prediction accuracy increases as the relatedness threshold becomes more permissive, mirroring the power gains documented above (see **Supplementary Tables 10 and 11** for full results). **Figure 3a** compares PGIs based on summary statistics from GRMA-S and GRMA-3^rd^ analyses in the UKB sample. Across all three phenotypes, the gain is substantial. **Figure 3b** shows that the pattern is qualitatively similar in within-family analyses conducted in the sample of All of Us^29^ siblings.

**Fig. 3.**
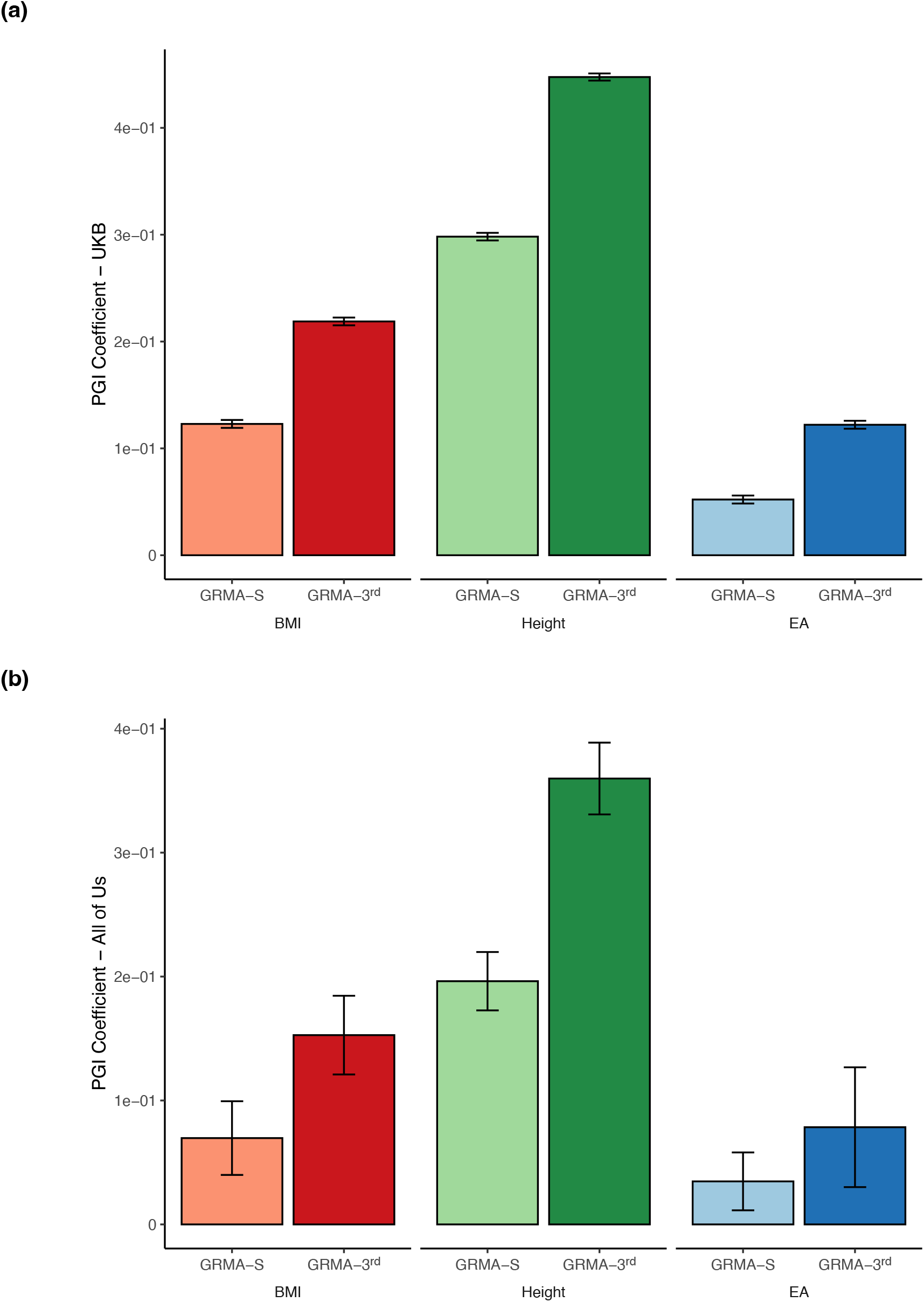
Predictive Power of GRMA Polygenic Indexes. Each panel shows the standardized regression coefficient of a phenotype on its corresponding PGI for BMI, Height, and Educational Attainment (EA). The *y*-axes show the magnitude of the coefficient. **(a)** Estimates in the *UKB*, using the (non-overlapping) “White British” Unrelated sample. **(b)** Estimates in *All of Us*, using the sample of siblings in a sibling-difference design. See **Online Methods** and **Supplementary Note** for additional details on samples and the methodology used to generate PGI weights.

Together, these findings demonstrate that GRMA yields substantially higher predictive power than sibling-only approaches. Indeed, compared to the predictive power of the most predictive PGIs constructed from any meta-analysis of family-based GWAS, Tan et al. (2025), our GRMA-3^rd^ PGIs have comparable regression coefficients for BMI (Tan et al.: 0.24 vs GRMA-3^rd^: 0.22) and larger predictive power for height (Tan et al.: 0.42 vs GRMA-3^rd^: 0.45) and educational attainment (Tan et al.: 0.08 vs GRMA-3^rd^: 0.12), despite being based only on UKB data.

This additional predictive power enables new insights. An example is the genetic correlation between educational attainment and BMI. Estimates based on population-GWAS summary statistics were negative^30,31^. Subsequent estimates that eliminate confounds by using family-based methods^12,32^ were imprecise and not statistically distinguishable from zero. Because GRMA is estimated in a diverse sample, standard LD scores are not applicable to estimate genetic correlation using LD score regression. However, we can show that the coefficient from a regression of (standardized) BMI onto the educational attainment PGI is a lower bound of the true genetic correlation (**Online Methods**). In the sibling-based All of Us sample, using the GRMA-3^rd^ educational attainment PGI to predict BMI, we obtain a regression coefficient estimate of -0.09 (SE = 0.03, *P* = 0.007). Our new evidence thus points once again to a negative genetic correlation, albeit possibly moderated in magnitude relative to the confounded, population-GWAS-based estimates.

#### Bias-Variance Trade-Off

A more direct approach to evaluating bias relies on the MMSE criterion, Equation (4), to determine which GRMA threshold is favored by the data given some weight parameter, *θ*. Using the estimated sampling variances and the squared bias estimate for GRMA-3^rd^ in **Supplementary Table 8**, we can calculate the range of *θ* where a researcher would prefer the GRMA-S results over the GRMA results at more permissive relatedness thresholds; we use *θ*^⋆^ to denote the threshold at which a researcher would be indifferent between the two sets of summary statistics. For both BMI and educational attainment, the threshold is *θ*^⋆^ = 0.01, implying that a researcher would only prefer the GRMA-S summary statistics if they put less than 1% of the weight on sampling variance. (Because the point estimate of the squared bias for GRMA-3^rd^ is negative for height, there is no value of *θ* for which the sibling-based estimates are preferred.)

This conclusion is robust even to conservative choices of the squared bias and sampling variance: under a “best-case” scenario for GRMA-S (using the *lower* bound of the 95%-confidence interval for 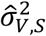) and a “worst-case” scenario for GRMA-3^rd^ (using the *upper* bounds of the 95%-confidence intervals for 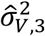 and 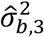), we find that the researcher would still require *θ* < 0.03 to prefer GRMA-S results for BMI and educational attainment and *θ* < 0.01 for height, still very distant from the conventional MSE criterion (*θ* = 0.5). While certain applications that use genetic data are particularly sensitive to bias (e.g., tests of polygenic selection^9,21^) and may require a lower *θ*, for ranges of *θ* that we view as defensible in most other cases, the MMSE criterion favors GRMA-3rd over GRMA-S. It is an open question whether this conclusion generalizes beyond these three phenotypes in UKB data, but the methodology is general: the GRMA framework provides a principled way to let the empirical evidence guide the choice of relatedness threshold.

### GWAS Benchmark

Next, we benchmark the GRMA results against summary statistics from a population-based GWAS conducted using standard protocols^33^. We conduct the GWAS in the UKB WB Unmatched sample (**Online Methods**). For comparability with the GRMA results, we restrict the GWAS to the same ∼830,000 HapMap3^34^ SNPs, use identical phenotype definitions to those in the main GRMA analyses, and conduct association analyses using a standard model that includes controls for sex, age, and principal components (**Online Methods**). While greater precision could be attained by including individuals from the GRMA sample, using a non-overlapping sample makes it straightforward to quantify the bias in the GWAS results relative to the GRMA-S results.

#### Genome-Wide-Mean Squared Bias

We first compare our benchmark GWAS to GRMA in terms of bias. We can reject the null of zero bias in the population GWAS summary statistics for all three phenotypes (BMI, *P* = 0.028; height, *P* = 4.8 × 10^-3^; educational attainment, *P* = 1.7 × 10^-6^). **Figure 4** compares the estimated genome-wide-mean squared bias in the benchmark GWAS to the GRMA-3^rd^ results (see **Supplementary Table 8** for complete results). For all three phenotypes, the estimated bias is substantially smaller in magnitude than in the GWAS benchmark and statistically indistinguishable from zero. These results suggest that GRMA-3^rd^ delivers substantial gains in power over GRMA-S without major sacrifices of robustness.

**Fig. 4.**
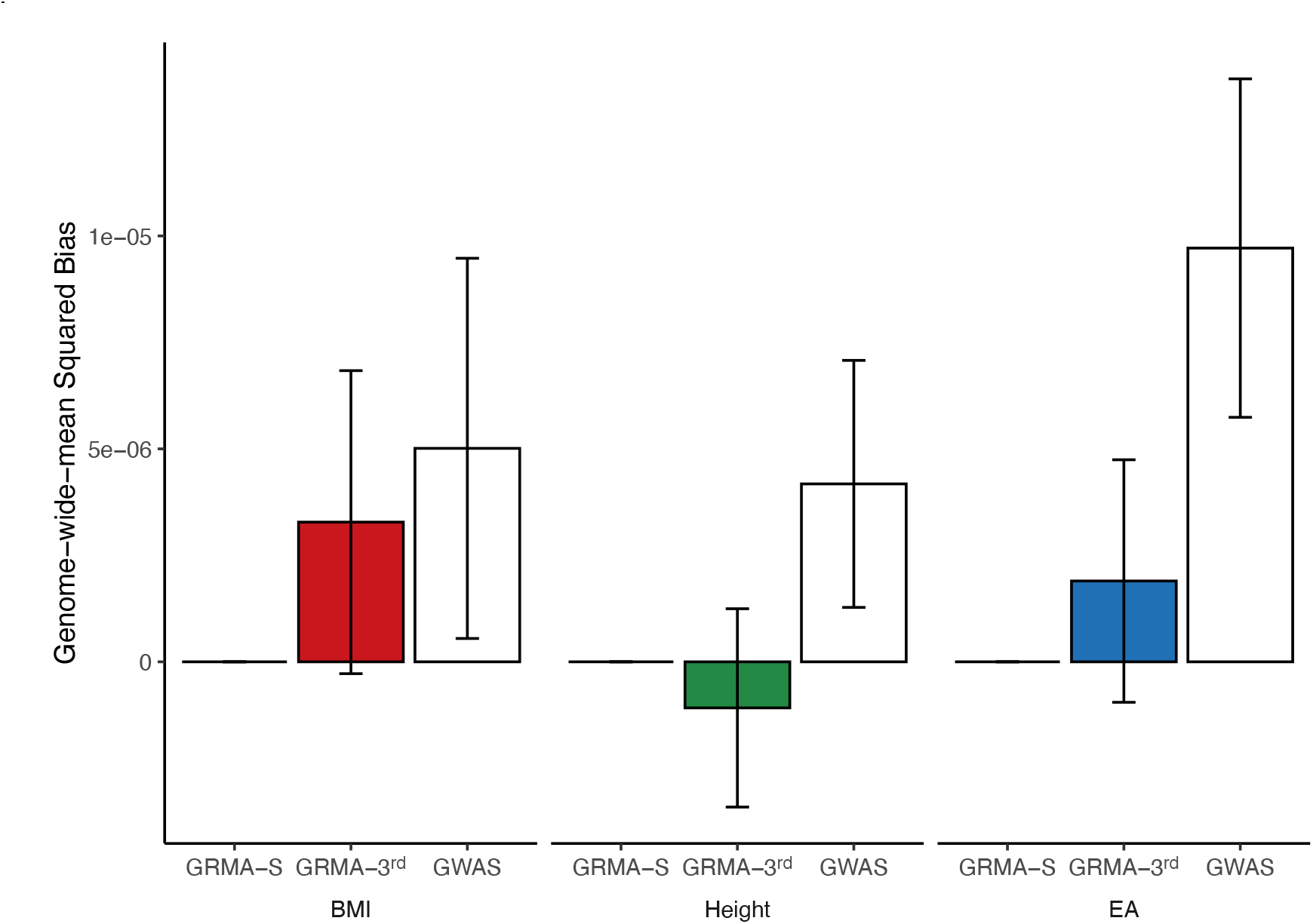
Genome-Wide-Mean Squared Bias. Genome-wide-mean squared bias in GRMA-S, GRMA-3^rd^, and Population GWAS for BMI, Height, and Educational Attainment (EA). The population GWAS was estimated in the sample of UKB “White British” Unrelated individuals. Bias is defined as expected deviation from the estimand of the GRMA-S estimates, which is why the bias for GRMA-S is, by definition, zero for all three phenotypes. See **Online Methods** for how the genome-wide-mean squared bias is estimated.

#### GRMA Provides Insights into Population GWAS Bias

GRMA-3^rd^ summary statistics remain vulnerable to some sources of bias, though generally to a lesser extent than population GWAS. Assuming that most 3^rd^-degree relatives in most datasets are first cousins and that these cousin pairs vastly outnumber the sibling pairs (as they do in the UKB), three biases are particularly relevant. First, because first cousins share one parental couple’s lineage (one of each cousin’s parents is a sibling of the other), GRMA-3rd removes about 12.5% of the bias due to parental genetic effects (“genetic nurture”), leaving the majority of this bias uncontrolled. Second, because first cousins share one set of grandparents, GRMA-3^rd^ reduces assortative-mating bias. The magnitude of the reduction is a complicated function of the heritability and sorting patterns. In a simple simulation of phenotypic assortment using estimates of heritability and parental phenotypic correlation from the literature^35,36^, GRMA-3^rd^ removed 48% of the assortative mating bias in BMI, 37% in educational attainment, and 50% for height (see **Online Methods**). Third, because cousins share at least half of their genetic ancestry, GRMA-3^rd^ can substantially reduce bias from population stratification. If there are many cousin pairs whose unrelated parents differ systematically in ancestry and allele frequency (e.g., due to recent admixture), some bias may remain. In summary, GRMA-3^rd^ should retain most of the bias contribution from parental genetic effects, much of the bias from assortative mating, and a small (but possibly positive) portion of the bias from population stratification.

Empirically, the estimated genome-wide-mean squared bias in the GRMA-3^rd^ results is substantially smaller than in the population GWAS: approximately 35% and 80% of the magnitude for BMI and educational attainment, respectively, and statistically indistinguishable from zero for both phenotypes. (The estimated genome-wide-mean squared bias for height is negative.)

Given the strong prior evidence of assortative mating for both height and educational attainment, the bias-reduction magnitudes of GRMA-3^rd^ relative to population GWAS are difficult to reconcile with large parental genetic effects (i.e., genetic nurture): if such effects were a dominant source of GWAS bias, GRMA-3^rd^ estimates would be expected to exhibit bias closer in magnitude to population GWAS. We therefore interpret these results as suggesting that, for these phenotypes in the UKB, parental genetic effects are unlikely to be a major contributor to genome-wide bias in population GWAS. This is consistent with the null estimates of parental genetic effects estimated using causal designs in the literature^37^. Instead, the majority of the bias in the population GWAS appears to be due to assortative mating and residual population stratification. While it is an open question whether this conclusion generalizes to other phenotypes and populations, GRMA provides a framework for assessing the relevance of these biases empirically.

## DISCUSSION

In this paper, we propose and evaluate GRMA, a new approach to genetic-association studies. GRMA improves on the now-standard study design, population GWAS, in two main ways: it can be implemented in ancestrally diverse samples and can reduce or largely eliminate bias. In our empirical application, we find that GRMA is computationally tractable and, even when applied only in UK Biobank (UKB), produces estimates of SNP associations that are more precise than those from the largest family-based GWAS meta-analysis. These properties translate into greater predictive power of PGIs constructed using summary statistics from GRMA.

The greater predictive power enables new insights. For example, existing family-based estimates of the genetic correlation between BMI and educational attainment have large confidence intervals. In contrast, the GRMA-3^rd^ educational-attainment PGI has a precisely estimated negative association with BMI. As another example, parental genetic effects are often cited as a primary source of bias in GWAS and PGI studies. However, for BMI, height, and educational attainment in the UKB, we find evidence that this is unlikely, with greater evidence of a major role of residual population stratification and assortative mating.

A potential limitation of these results is that, although we do not detect any remaining bias in our GRMA summary statistics, our simulation results imply that some bias from assortative mating may remain. This means, for instance, that the genetic correlation we find between BMI and educational attainment may partly be driven by cross-trait assortative mating^38^. However, even considering the upper bound of our estimate of the genome-wide-mean squared bias, we find that the bias must be small relative to the bias in a population GWAS for height and educational attainment.

In addition to its other advantages, GRMA offers a way of conducting genetic-association research in large samples without the need to assign individuals to discrete (e.g., continental) genetic ancestry groups. In so doing, GRMA avoids the wasteful practice of discarding data on individuals with underrepresented or unclassifiable ancestries and does not contribute to reifying racial categories^13^.

While GRMA is most closely related to family-based GWAS, it also has similarities with structured-association approaches^39,40^. These studies identify groups based on genetic relatedness (often with groups much smaller than continental ancestry^41^) and then control for each person’s group membership (allowing partial membership in the case of admixture). In contrast, GRMA assigns individuals to unique, potentially overlapping groups based on a person’s genetically inferred pedigree. Thus, it relies on within-group comparisons of people that are generally more closely related than groups from structured association studies.

GRMA summary statistics can be used in place of GWAS summary statistics in downstream applications, e.g., for heritability partitioning^42,43^, gene identification and fine-mapping^44–47^, and transcriptomics^48^. However, because GRMA is robust in diverse-ancestry samples, the linkage-disequilibrium patterns that underlie its estimates may differ from that of single-ancestry linkage-disequilibrium panels used in many methods. Therefore, appropriate panels will need to be developed for summary statistics from diverse-ancestry GRMA studies.

The GRMA approach could be extended to the estimation of other genetic-association parameters. For example, PGI studies have become a workhorse in research using genetic data in psychology, economics, sociology, and epidemiology^49,50^, but causal analyses have been difficult to conduct because genotyped sibling or trio data is scarce. However, demeaning PGIs within, say, third-degree relative groups, could greatly increase statistical power while introducing little bias in many circumstances.

Beyond its specifics, GRMA illustrates the more general point that extended relatives are an untapped resource. As biobanks around the world collect data in ever larger samples, extended relatives are, by chance, increasingly included. Taking advantage of these relationships could greatly accelerate genetics research.

## DATA AVAILABILITY AND ACCESSION CODES

Data from the UK Biobank and All of Us, which were used in this study, are available to researchers upon application.

Upon publication, summary statistics produced as part of this paper can be downloaded from the Social Science Genetics Association Consortium Data Portal, www.thessgac.com.

## CODE AVAILABILITY

Upon publication, all scripts and code used for data analysis and producing the figures and tables will be made available in a publicly available GitHub repository.

## URLs

GRMA: https://github.com/JonJala/grma

Social Science Genetic Association Consortium (SSGAC) website: http://www.thessgac.org/#!data/kuzq8.

Plink: https://www.cog-genomics.org/plink/

Kinship-based INference for Gwas (KING): https://www.kingrelatedness.com

snipar: https://snipar.readthedocs.io/en/latest/guide.html

## ACKNOWLEDGEMENTS

We gratefully acknowledge the research participants of the UK Biobank and All of Us cohorts, without whom this research would not be possible. This research has been conducted using the UK Biobank resource under Application Number 11425 and data from the All of Us Research Program’s Registered/Controlled Tier dataset. We also thank the National Institutes of Health’s All of Us Research Program for making available the participant data examined in this study. The study was supported by Open Philanthropy and the National Institute on Aging/National Institutes of Health through grants R01-AG081518 to P.T., R24-AG065184 and R01-AG042568 to D.J.B., and R01-AG083379 to A.S.Y. We also thank Michael Edge, Peter Visscher, Aysu Okbay, Matthew Howell, Robel Alemu, Loic Yengo, other members of the Social Science Genetics Association Consortium community, and the participants at the BGA, IGSS, and TAGC conferences for their invaluable feedback on this research.

## AUTHOR CONTRIBUTIONS

P.T., D.C., and D.B. conceived and designed the study.

D.J. was the lead analyst, designing the software, cleaning the data, and carrying out the other analyses found in this paper.

T.G. led the analyses at early stages of this project and carried out background research that informed the ultimate design of the GRMA study method.

J.J. supported the design of the GRMA software.

J.S. provided research support and helped design the figures and tables of the paper.

A.Y. aided in study design and in the comparison of GRMA to the snipar method.

P.T., D.J., and D.C. wrote the first draft of this manuscript and its supplement.

All authors reviewed and provided critical feedback on the final draft.

## COMPETING INTERESTS

A.Y. is an advisor to and holds equity in Herasight, Inc. The remaining authors declare no competing interests.

## ONLINE METHODS

This article is accompanied by a **Supplementary Note** with further details.

### Theoretical Framework

For each individual *i* = 1,2, …, *N* and SNP *k* = 1,2, . . ., *K*, let *y*_*i*_ denote the phenotype, *x*_*ik*_ denote the genotype, and *f*_*ik*_ the mean parental genotype, all demeaned at the population level. Throughout, we use ordinary font for scalars (e.g., *y*_*i*_, *x*_*ik*_) and boldface font to denote vectors or matrices.

The population-level regression model for SNP *k* is:

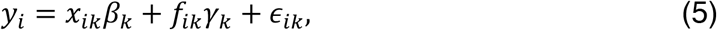

Ordinary least squares (OLS) regression of *y*_*i*_ onto *x*_*ik*_ controlling for *f*_*ik*_ yields unbiased estimates of *β*_*k*_, because due to Mendelian inheritance of alleles, *x*_*ik*_ is uncorrelated with *ϵ*_*i*_ *conditional* on *f*_*i*_. That is, 𝔼(*x*_*ik*_*ϵ*_*ik*_|*f*_*ik*_) = 0 for every *k* = 1,2, . . ., *K*. For notational compactness, we sometimes omit the SNP subscripts if it is clear from context that the regression is at the level of an arbitrary SNP.

A fundamental challenge for applied work is that empirical analogs of Eq. (5) are usually not feasible because the mean parental genotype is almost never observed, and outside of special designs with limited power, such as sibling regressions, cannot be differenced out.

### Relatedness Groups

In Genomic-Relatedness-Matched Association (GRMA) studies, the empirical challenge highlighted in the previous section is addressed by exploiting variation within *relatedness groups*: sets of individuals who are “sufficiently” genetically related, as defined according to some criterion. Intuitively, estimating each genetic association *within* relatedness groups reduces bias by controlling for relatedness-group-level genetic background, since members of the same group draw their alleles from more similar parental distributions than unrelated individuals. While this is not in general guaranteed to eliminate bias, it often provides a more robust alternative to population-based GWAS.

GRMA relatedness groups are constructed in two steps.

#### Step 1: Relationship Inference

Starting from a quality-controlled sample of *N* individuals, all pairwise relationships are inferred. We use the KING software^26^ to estimate two relatedness parameters per pair (*i, j*): a kinship coefficient 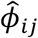 and a probability of zero identity-by-descent (IBD) sharing, 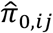. Relationship types are then assigned by comparing 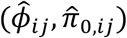 to the theoretical ranges implied by quantitative-genetic theory (Table 1 of ref. ^26^). By construction, the regions in the (*ϕ, π*_0_) space corresponding to the relationship types we study – siblings, parent-offspring, 2^nd^-degree, and so on – are mutually exclusive. It follows that whenever a relationship can be assigned, it is unique. Pairs that fail to meet the inference criteria at the most permissive relatedness threshold specified are classified as unrelated.

#### Step 2: Group Formation

Our algorithm forms groups hierarchically by relationship type, beginning with the closest relatives (full siblings) and proceeding stepwise to more distant categories, up to the maximum degree specified by the user (up to 3^rd^-degree relatives). The algorithm cycles through individuals *i* = 1,2, . . ., *N*, constructing one relatedness group for each individual.

For a given individual *i*, the procedure is:

1. Full siblings. The algorithm checks whether any full siblings exist in the full *N* sample. If so, it adds **all such siblings** to ℛ_*i*_. At that point, the group is finalized at the sibling level, and no more distant relatives are considered for ℛ_*i*_.
2. Parent–offspring. If no siblings are found, the algorithm checks for parents or offspring, again in the full *N* sample. If at least one is identified, **all such first-degree relatives** are added, and the group is then finalized.
3. Second-degree relatives. If no first-degree relatives are found in Steps 1 and 2, the search proceeds to second-degree relatives, again in the full sample, adding **all such relatives** if any exist, and then finalizing the group.
4. Third-degree relatives. If no second-degree relatives are found in Steps 1 through 3, the search proceeds to third-degree relatives, again in the full sample, adding **all such relatives** if any exist, and then finalizing the group.
5. Unmatched individuals. If no relatives were identified in the sample in Steps 1 through 4, the individual is labeled as Unmatched.

We highlight some properties of the algorithm.

For each individual, the procedure begins anew. For example, when forming *i*’s relatedness group, if *i* has siblings, and *j* is *i*’s cousin, then *j* will not be assigned to ℛ_*i*_, since ℛ_*i*_ was formed and closed in Step 1. But when forming *j*’s relatedness group, if *j* is not matched to any relatives in Steps 1 through 3, then *i will* be assigned to ℛ_*j*_ in Step 4.

It follows that membership in a relatedness group need not be either symmetric or transitive. To illustrate, consider a GRMA analysis of four individuals – *A, B, C*, and *D* – whose pedigree chart is shown in **Fig 1a**. Here, *A* and *B* are full siblings, and *C* has no full siblings but is the first cousin of *A, B* and *D*. Given these relationships, the algorithm will produce the following relatedness groups: ℛ_*A*_ = {*A, B*}, ℛ_*B*_ = {*A, B*}, ℛ_*C*_ = {*A, B, C, D*}, and ℛ_*D*_ = {*C, D*}. In this scenario, symmetry is violated since *A* is in ℛ_*C*_ even though *C* is not in ℛ_*A*_. Transitivity is also violated since *C* is in ℛ_*D*_ and *A* is in ℛ_*C*_, but *A* is not in ℛ_*D*_.

### The Residualization Matrix and Within-Group Model

Let **Y, X, F**, and **ϵ** denote the length-*N* column vectors corresponding to the realized values of *y*_*i*_, *x*_*ik*_, *f*_*i*_, and *ϵ*_*i*_. Omitting the SNP subscript *k*, we can then write the empirical analog of Equation (1) compactly as:

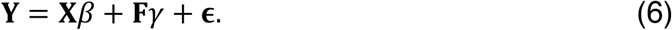

Let ℛ_*i*_ ⊆ {1,2, . . ., *N*_*i*_} denote the set of members of *i*’s relatedness group. Denote its size by *N*_*i*_ = |ℛ_*i*_|. To implement the within-group association analyses, define the *N* × *N* matrix **R** whose diagonal entries are **R**_*ii*_ = (1 − 1/*N*_*i*_) and whose off-diagonal entries are, for every *i* ≠ *j*:

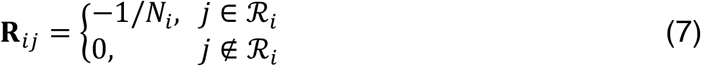

Premultiplying any length-*N* vector **v** by **R** transforms each element to a deviation from its relatedness-group mean:

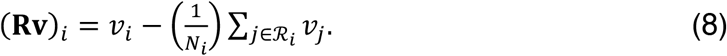

In what follows, we use tildes to indicate a variable has been demeaned at the relatedness-group level, so that 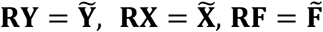 and 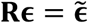.

Applying the transformation to the data yields the within-group model:

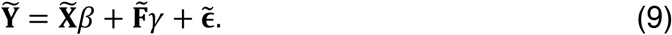

### GRMA Estimator

The GRMA estimator for a given SNP is then OLS applied to the regression of 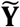 on 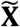:

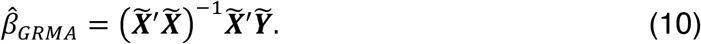

The vector 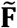 is unobserved so it is not included in the regression. We therefore define the composite residual

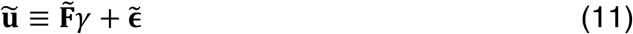

and write the model as 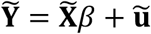. This decomposition highlights the two potential sources of bias, as discussed in the main text:

i. 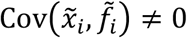, which arises when there is residual stratification even within relatedness groups. For example, this may occur when the unrelated parents of a pair of cousins have different genetic ancestries, as might be the case with recent admixture.
ii. 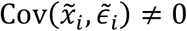, which arises when an individual’s outcome is impacted by the genotypes of their relatives, a phenomenon called interpersonal genetic effects or indirect genetic effects. For example, if *i* and *j* are siblings and *j*’s genotype *x*_*j*_ affects *i*’s phenotype (i.e., sibling genetic effects), then *x*_*j*_ is part of *i*’s residual *ϵ*_*i*_. Therefore 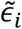 and 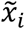 are both functions of *x*_*j*_ and are thus correlated. This correlation produces bias in the GRMA regression.

A special case of interest arises when relatedness groups consist exclusively of full siblings. In this setting, for any *j* ∈ ℛ_*i*_, we must have *f*_*i*_ = *f*_*j*_, so that relatedness-group demeaning eliminates the unobserved parental midpoint exactly. The only remaining potential source of bias in this special case is due to sibling genetic effects, which are present if one of the unobserved factors *ϵ*_*i*_ influencing *y*_*i*_ is sibling *j*’s genotype *x*_*j*_, generating bias (ii) above.

### GRMA Standard Errors

In the **Supplementary Note**, we derive the following closed-form expression for the sampling variance of 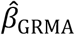:

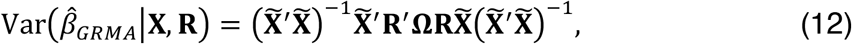

where **Ω** = Var(**F***γ* + **ϵ**). While 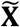 and **R** are observed, **Ω** is unknown. We obtain an estimate of this matrix, 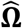, assuming that *β* is small (such that **F***γ* + **ϵ** ≈ **Y**), that the covariance of **F**_*i*_*γ* + **ϵ**_*i*_ between any pair of individuals depends only on their type of relatedness (e.g., full-siblings, parent/offspring, etc.), and that the covariance of **F**_*i*_*γ* + **ϵ**_*i*_ between pairs of individuals more distant than 3^rd^-degree relatives is zero. The feasible variance estimator therefore plugs 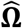 into the expression of the sampling variance, obtaining

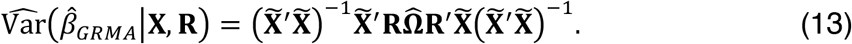

For a full derivation and further discussion, see **Supplementary Note**.

### Simulation Evidence on Standard Error Calibration

We assess whether the GRMA standard errors are correctly calibrated in a simulation. Specifically, we simulate an extended pedigree such that 4 people’s closest relative is a full sibling, 1 person’s closest relative is a Parent/Offspring, 2 people’s closest relative is a 2^nd^ degree, and 7 people’s closest relative is a 3^rd^ degree. This pedigree is repeated 10,000 times to approximate sample sizes for each relatedness threshold in the UK Biobank (UKB) data. (See **Table 1**.) In this simulation, we generate genotypes for 200 SNPs in linkage equilibrium, one of which is null and 199 are causal. The effect sizes for causal SNPs are drawn from an independent, identically distributed normal distribution and the residual is drawn from a normal distribution with variance fixed such that the heritability of the phenotype is 40%. We then use GRMA to estimate the effect of the null SNP using full-sibling, parent/offspring, 2^nd^-degree, and 3^rd^-degree thresholds. The same genotypes but different effect sizes and residuals are used over 10,000 replications. **Supplementary Figure 1** contains quantile-quantile (QQ) plots of the p-values for each relatedness threshold. Across 10,000 replications, the rejection rate at each significance level is consistently very close to expectations, suggesting that GRMA standard errors are well calibrated for correct inference in settings similar to those in our application (see **Supplementary Note** for additional details on the simulations).

### Bias-Variance Tradeoff

To formalize the tradeoff between bias and variance that arises when selecting a relatedness threshold, we define a modified mean-squared error criterion. For each SNP *k* = 1,2, . ., *K*, define

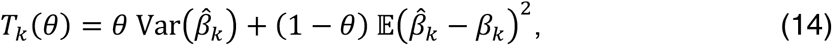

where we use 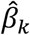 as a shorthand for 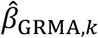. The parameter *θ* ∈ [0,1] specifies the weight placed on the variance term, 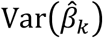, relative to the bias term, 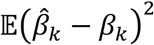, with *θ* = 0.5 corresponding to the usual mean-squared error criterion. The modified mean-squared error criterion aggregates this per-SNP criterion across all *K* SNPs:

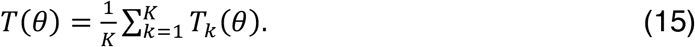

In most empirical settings, the bias component of mean-squared error cannot be measured in practice since the true population parameter is unknown. An attractive feature of GRMA is that in any dataset containing siblings, we can estimate the genome-wide average bias relative to the estimand of the sibling-based estimator.

#### Estimation of Genome-Wide Sampling Variance, 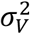

Our estimator of the average genome-wide sampling variance, 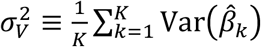, is:

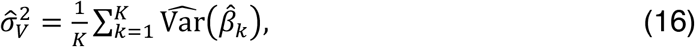

where we use the squared estimated standard errors for 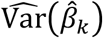.

#### Estimation of Genome-Wide Squared Bias, 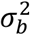

For the estimate of the average genome-wide squared bias, 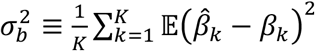, in the **Supplementary Note**, we derive the following expression for 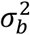:

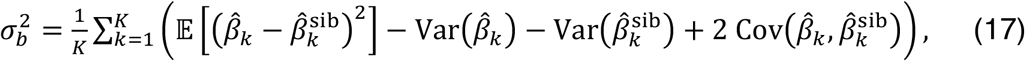

and show that

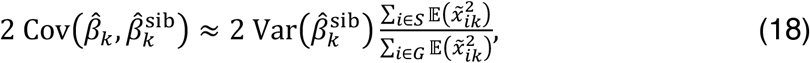

where *S* and *G* are the sets of individuals in the sibling sample (i.e., those with at least one sibling in the sample) and the GRMA sample (i.e., those with at least one other person in the relatedness group responding to the relatedness threshold used to obtain 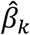), respectively.

Together, these conditions suggest the following feasible estimator of the genome-wide average squared bias:

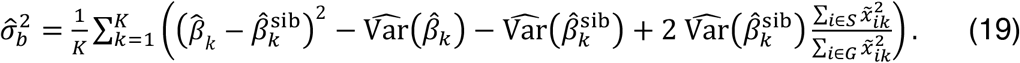

For a full derivation and additional analyses of the properties of this estimator, see **Supplementary Note**, which also validates that it has intuitive properties in two special cases. First, when 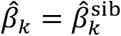 (i.e., the GRMA study is based on a sibling threshold), the bias estimate is zero by construction. Second, in the limiting case of an infinite sample, the feasible estimator of squared bias converges to the true population squared bias.

#### Block Jackknife for Inference

To obtain standard errors for 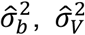, and 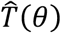, we implement a block-jackknife procedure over SNPs that divides the genome into 200 contiguous blocks. For each block *c* = 1,2, . . .,200, we recompute the estimator with the block left out. For each estimator, 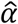, the jackknife variance for the genome-wide squared bias is then estimated as:

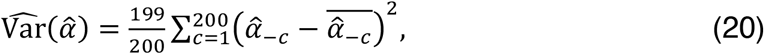

where 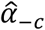 is the recomputed estimator with block *c* omitted and 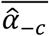 is the mean of the 200 recomputed estimators with one block omitted in each. The jackknife procedure takes linkage disequilibrium into account by treating nearby SNPs as a block rather than as independent observations.

### Empirical Applications

Our empirical applications are designed to evaluate the performance of GRMA across relatedness thresholds and relative to two established methodological approaches, population GWAS and sibling GWAS. The analyses are all conducted in UKB and consider three phenotypes: body-mass index (BMI), height, and educational attainment. In addition, we evaluate the predictive performance of polygenic indices constructed from GRMA summary statistics in a sample of siblings in the All of Us cohort.

### SNP-Level Filters

Our analyses are restricted to HapMap3 SNPs passing standard variant-level quality controls, including minor allele frequency (MAF) above 0.01, INFO score above 0.99, missingness per marker < 5%, and Hardy-Weinberg Equilibrium (HWE) test p-value above 1e-6. In a diverse sample, SNPs can fail the HWE test both due to population stratification and poor genotype calling. Because GRMA is robust to population stratification, we only wish to drop SNPs that fail due to the poor genotyping. Therefore, we apply the HWE p-value test only within the sample defined as “White British” by the UKB. Applying these filters leaves *K* = 828,208 SNPs (see **Supplementary Table 3**).

### Subject-Level Filters

To arrive at our estimation sample, we begin with the full sample of genotyped UKB participants (excluding any participants who had elected to withdraw at the time of our analyses). We apply standard subject-level quality control filters, including dropping individuals with sex chromosome aneuploidy, excess heterozygosity, excess relatives, ambiguous sex, and missingness per individual per chromosome > 5%. After applying these filters, the analysis sample includes *N* = 483,712 individuals, of whom 407,759 are in the “White British” subsample defined by the UKB^17^. GRMA does not require filters on ancestry that are typically required by population GWAS. This allows for greater utilization of the sample and showcases GRMA’s ability to be performed in a diverse sample.

### Phenotypes

We use BMI and height measurements collected from participants at their UKB baseline assessment visit, which included measurement of weight and standing height. BMI is defined as weight (in kilograms) over squared height (measured in meters), and height is measured in centimeters. Educational attainment was measured in years of schooling, imputed from responses to survey questions in UKB about highest educational qualification. We follow prior work by assigning a year-of-schooling equivalent to each qualification, using the International Standard Classification of Education (ISCED) mapping designed to harmonize educational measurement across countries^5^. Descriptive statistics for all three outcome variables are shown in **Supplementary Table 1**. To facilitate the comparison of GRMA results across phenotypes, for each relatedness threshold, we standardize each phenotype by sex among the set of individuals who have at least one person in their relatedness group. For example, when conducting a GRMA study at a 2^nd^-degree threshold, the phenotype is standardized by sex among those who have at least one 2^nd^-degree relative or closer in the full sample.

### Mean *χ*^**2**^ Statistic

One way we characterize differences between GRMA study results based on different relatedness thresholds is the genome-wide mean *χ*^2^. We use GRMA-S, GRMA-PO, GRMA-2^nd^, and GRMA-3^rd^ to denote GRMA studies with relatedness thresholds at the sibling, parent-offspring, 2^nd^-degree relative, and 3^rd^-degree relative thresholds, respectively. For each set of GRMA summary statistics, one per relatedness threshold, we compute

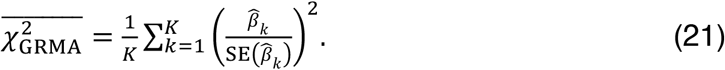

Results for each of the GRMA analyses for 4 relatedness thresholds and 3 phenotypes are shown in **Supplementary Table 7**.

### GRMA-S-Equivalent Sibling Effective Sample Size

We interpret the mean *χ*^2^ statistic results by evaluating how much larger a GRMA-S study (or equivalently a sibling-based GWAS) would have to be to achieve the same mean *χ*^2^ statistic that is obtained in the GRMA summary statistics for more permissive thresholds. Following the approach outlined in Turley et al.^51^, we can show that one minus the mean *χ*^2^ statistic of sibling GWAS summary statistics is proportional to the sibling effective sample size, defined as *N*_eff_ ≡ *N* − |*N*_*D*_|, where *N* is the total number of siblings and |*N*_*D*_| is the number of families in the sample. Therefore, the relationship between the GRMA- and sibling-based mean *χ*^2^ statistic 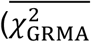 and 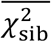, respectively), the sibling effective sample size (*N*_eff_), and the GRMA-equivalent effective sample size (*N*_GRMA-equiv_) is

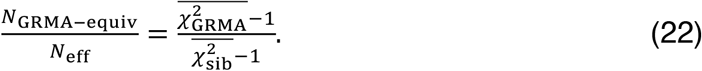

Under the assumption that new families are drawn from the same population distribution as the observed sample (which includes the assumption that the distribution of family size is the same), this ratio is how much the sibling sample size would need to multiplied to produce the same mean *χ*^2^ statistic that is obtained in the GRMA summary statistics.

### Benchmark Analyses

We compare GRMA estimates to two benchmark approaches: population GWAS and family-based snipar summary statistics. As mentioned in the discussion of the bias-variance tradeoff, we calculate the genome-wide-mean squared bias relative to the GRMA-S estimand across the GRMA, population GWAS, and snipar summary statistics. As proven in the **Supplementary Note**, the genome-wide-mean squared bias is independent of sample size, which facilitates comparability between GRMA and population GWAS.

#### Population-Level GWAS

In order to produce a benchmark population GWAS to compare to GRMA, we run a GWAS in individuals of White British ancestry in the UKB. To ensure no overlap with GRMA estimation samples, we use White British individuals who are Unmatched in the relatedness group formation (i.e., those who do not have any relative as close as a 3^rd^ degree relation in the sample). (See **Table 1**.) In addition to the QC procedure, we apply in the sections **SNP-Level Filters** and **Subject-Level Filters** above, we also filter out individuals who have a PC value that is greater than 5 SDs from the mean on any of the first 10 PCs. We use PC values computed and distributed by the UKB. We standardize each phenotype within sex and use the first 10 PCs as controls. This results in a sample size of approximately 275,000 individuals. Full counts of the sample size per phenotype are detailed in **Table 1**.

In the population GWAS, we use the same three phenotypes (BMI, height, and educational attainment), constructed as described in the above section **Phenotypes**.

For each measured SNP, the association model used in the population GWAS is:

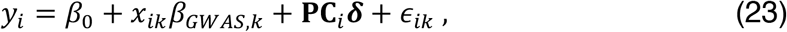

where *x*_*ik*_ is the best-guess allele dosage of SNP *k*. Here, using the best-guess allele dosage should be almost equivalent to using the full probabilistic dosages because of our strict INFO score filter (greater than 0.99), which results in keeping only SNPs with high imputation accuracy. **PC**_*i*_ is a vector of *i*’s values for the first 10 principal components of the variance-covariance matrix of the genotypic data. This analysis was carried out in plink 2.0 which estimates effects using OLS for continuous phenotypes as in our application. All GWAS were run on the same harmonized set of ∼800K high-quality HapMap3 SNPs used for GRMA to keep SNP coverage comparable.

#### Snipar-Robust and snipar-Unified

In addition to comparing our results to a standard population GWAS, we also evaluated how GRMA performed compared to other family-based methods that improve bias and precision relative to population GWAS: snipar-Robust and snipar-Unified^23^.

For each phenotype, we compare GRMA results to publicly available, published summary statistics from snipar-Robust and Unified analyses. These summary statistics were produced by running snipar on phenotypes that were standardized by sex in the UKB, making them comparable to the phenotypes and sample used for our GRMA results. Snipar-Robust was run on a sample of size *N* = 51,875 individuals without ancestry restrictions. Snipar-Unified was run on a sample of *N* = 408,254 individuals classified as having “White British” ancestry.

Details on snipar-Robust and snipar-Unified can be found in the paper that presented the methods^23^. To briefly summarize, snipar-Robust increases power to estimate genetic effects using within-family variation in genotypes even when both parents aren’t observed. It can obtain estimates that are more powerful than sibling-based estimates that are robust to most sources of confounding, even in diverse samples. Snipar-Unified builds on the imputation methodology developed in the original snipar paper^52^ by including samples without genotyped first-degree relatives through linear imputation. While this can increase power, it can be biased when there is strong population structure and/or admixture. In relatively homogeneous samples, however, the bias is negligible. For this reason, the unified estimator is applied in the White British subsample of the UKB.

We subset both GRMA and snipar results to the intersection of SNPs across all summary statistics files to keep SNP coverage and sampling variance comparable, resulting in ∼141,000 SNPs. Estimates of the sampling variance of the GRMA and snipar approaches are shown in **Supplementary Table 9**.

### Comparison Metrics

We consider two metrics to compare the GRMA results to the GWAS and snipar results.

#### Genome-Wide-Mean Squared Bias

Using the same procedure described in the above section, **Estimation of Genome-Wide-Mean Squared Bias**, we can estimate the bias of the GWAS estimates relative to the GRMA-S estimand because the covariance term of Equation (17) is zero. This is because there is no sample overlap between the GRMA and GWAS samples. The bias of the snipar-Robust estimates is assumed to be zero. We are unable to estimate the bias of the snipar-Unified estimates because we do not know the covariance between the snipar estimation error and the GRMA-S estimation error. GWAS mean squared bias estimates for the set of 828,208 SNPs tested by GRMA are reported in ***Supplementary Table 8***.

#### Genome-Wide-Mean Sampling Variance

We estimate the genome-wide-mean sampling variance for snipar-robust and snipar-unified using the same procedure described in the above section, **Estimation of Genome-Wide-Mean Sampling Variance**. The comparison of snipar, population GWAS, and GRMA estimates are reported in ***Supplementary Tables 8 and 9***. However, we highlight that a comparison of the genome-wide-mean sampling variance population GWAS with that of snipar and GRMA is not necessarily fair. In practice, population GWAS often include meta-analyses of many cohorts that have few relatives in them. Therefore, considering our population GWAS of just the unrelated White British subsample of the UKB ignores the large amount of data that might be used if one were performing a population GWAS.

For both estimators of the genome-wide-mean squared bias and mean sampling variance, we produce standard errors of the estimates by using a block-jackknife procedure as detailed in *Block Jackknife for Inference* above.

### Polygenic Prediction

We evaluated the predictive performance of polygenic indices (PGIs) constructed from GRMA summary statistics at each relatedness threshold. PGI weights were generated using SBayesR^28^ to correct for LD using a Bayesian multiple regression model with a mixture of 4 normal distributions for the prior on the SNPs’ effect sizes. Prior mixture proportions were set to 0.95, 0.02, 0.02, and 0.01 with the variance of the distribution of genetic effects set at 0, 0.01, 0.1, and 1, respectively. We used pre-computed sparse and shrunk LD matrices for HapMap3 variants from Lloyd-Jones et al.^28^ Each Markov chain was run for 10,000 iterations with a burn-in of 2000. Because the GRMA results are based on the diverse UKB sample and not a sample with matched ancestry to the reference panel, the predictive accuracy of the resulting PGIs may be reduced. However, the predictive accuracy of all PGIs would be reduced comparably within a phenotype, making the comparison reasonable.

Prediction accuracy was assessed in two hold-out samples:

1. UKB Unmatched “White British” individuals: out-of-sample prediction of BMI, height, and educational attainment in UKB individuals not included in any GRMA estimation samples.
2. All of Us siblings: within-family prediction of BMI, height, and educational attainment.

#### UKB Unmatched “White British”

We carry out our prediction analysis in the same sample of Unmatched White British UKB individuals that we use for our benchmark population GWAS. Because these individuals are by construction non-overlapping with and not closely related to the GRMA sample, they constitute a valid prediction sample. We use Plink to produce PGIs using the SBayesR-adjusted GRMA summary statistics and the UKB genotypes. We have 12 PGIs corresponding to the 4 GRMA relatedness thresholds and 3 phenotypes. We standardize each PGI within the prediction sample.

#### UKB Singleton Prediction Model

We estimate a prediction model of the form

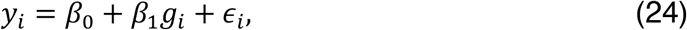

where *y*_*i*_ is the phenotype and *g*_*i*_ is the PGI, both standardized within the prediction sample. We define prediction accuracy using 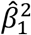—which is equivalent to the *R*^2^ of the regression because the phenotype and PGI are standardized—and find results that align closely with theoretical predictions. Results for the UKB prediction analysis are detailed in **Supplementary Table 10**.

#### All of Us siblings

All of Us provides a table of pairwise kinship values for all individuals in the sample. We use this table to determine the set of individuals who have at least one 1^st^-degree relative in the sample, and we run KING on this subset to determine the set of individuals who have at least one sibling in the sample. We do not restrict the prediction sample based on ancestry. This results in ∼12,800 individuals with at least one sibling in the sample.

#### Phenotypes

All of Us collects physical measurements such as weight and height from in-person clinical visits, self-reported questionnaires, and EHR extraction. We use the BMI (measured in *kg*/*m*^2^) and height variables (measured in centimeters) provided by All of Us. Following previous studies^5^, we construct an educational attainment phenotype by mapping responses to the question ‘What is the highest grade or year of school you completed?’ to International Standard Classification of Education levels. Further details regarding this mapping are in the **Supplementary Note**. We standardize all phenotypes by sex at birth among the set of individuals in the sample with at least one sibling. Summary statistics regarding phenotypes are detailed in **Supplementary Table 1**.

#### PGI Construction

We use plink to construct PGIs using All of Us sibling genotypes and the SBayesR-adjusted GRMA summary statistics from the UKB. Thus, we have 12 PGIs corresponding to the 4 GRMA relatedness thresholds and 3 phenotypes. We standardize each PGI.

#### Prediction Model

To estimate the predictive power of each PGI, we first demean the phenotypes and PGIs within each family, producing demeaned phenotypes, 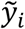, and PGIs 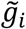. Next, we drop one sibling per family. We then estimate

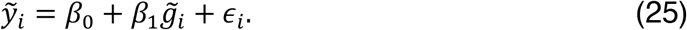

We use 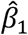 to denote the estimate of *β*_1_ from this regression. Finally, to account for correlation within each family, SEs are clustered at the sibling group level. We define prediction accuracy using 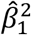. Because the phenotype, *y*_*i*_, and PGI, *g*_*i*_, are standardized, 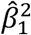 is an estimate of the fraction of variance in the phenotype that is due to causal variation in the PGI. To see why, note that

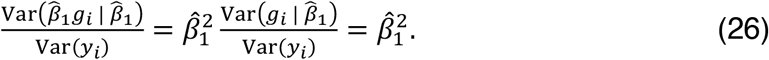

#### Theoretical Prediction

Using results from Daetwyler et al.^53^ we can calculate theoretical predictions of our PGI predictive power from GRMA-S estimates in the sibling sample of All of Us:

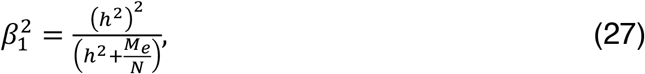

where *h*^2^ is SNP heritability. *M*_*e*_ is the effective number of independent SNPs which we assume to be 60,000,^54^ and *N* is GRMA effective sample size. For GRMA-S summary statistics, the effective sample size is the total number of siblings minus the number of families. In **Supplementary Table 11**, we show theoretical predictions of *β*_1_ for BMI, height, and educational attainment. Using estimates of SNP heritability from Tan et al.,^32^ we find that these theoretical predictions are very close to our estimates of predictive power 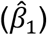.

### Predicting BMI with the Educational Attainment PGI

To test for a genetic correlation between educational attainment and BMI, we use the BMI phenotype and the educational attainment PGI in the sibling sample in the *All of Us* data described above. We then regress BMI onto the educational attainment PGI and report the regression coefficient. This coefficient is an estimate of the genetic correlation between BMI and educational attainment attenuated toward zero. To see why, first we express BMI as

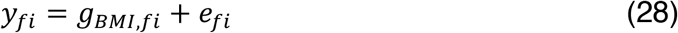

where *y*_*fi*_ is the standardized BMI measure for individual *i* from family *f, g*_*BMI,fi*_ is that individual’s genetic factor for BMI (such that 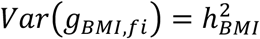 is the heritability of BMI), and *e*_*fi*_ is the residual. Next, we express the standardized educational attainment PGI as

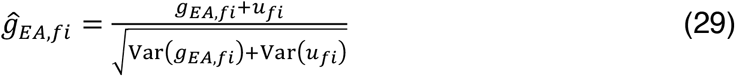

where *g*_*EA,fi*_ is the true genetic factor for educational attainment (such that 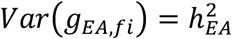 is the heritability of educational attainment), and *u*_*fi*_ is the estimation error in the PGI due to sampling variance in the PGI weights. (For example, if the PGI weights were estimated in an infinite sample, then there would be no estimation error, *u*_*fi*_ = 0, and *ĝ*_*EA,fi*_ = *g*_*EA,fi*_.) We allow for *g*_*BMI,fi*_ and *e*_*fi*_ to be correlated and for *g*_*EA,fi*_ and *e*_*fi*_ to be correlated, but we assume that the sibling difference *e*_*f*1_ − *e*_*f*2_ is uncorrelated with both *g*_*BMI,f*1_ − *g*_*BMI,f*2_ and *g*_*EA,f*1_ − *g*_*EA,f*2_. Also, because *u*_*fi*_ is due to sampling variance in the PGI weights, we assume it is uncorrelated with all other variables in the model, which is a reasonable assumption as long as the educational attainment GWAS sample and the prediction sample don’t overlap^49^. Finally, for simplicity in this illustration, we assume random mating, which implies Var(*g*_*BMI,f*1_ − *g*_*BMI,f*2_) = Var (*g*_*BMI,fi*_ ), Var(*g*_*EA,f*1_ − *g*_*EA,f*2_) = Var(*g*_*EA,fi*_), Var(*ĝ*_*EA,f*1_ − *ĝ*_*EA,f*2_) = Var(*ĝ*_*EA,fi*_), and Cov(*g*_*BMI,f*1_ − *g*_*BMI,f*2_, *g*_*EA,f*1_ − *g*_*EA,f*2_) = Cov(*g*_*BMI,fi*_, *g*_*EA,fi*_).

Under these assumptions, we can express the coefficient from regressing BMI onto the educational attainment PGI in a sibling difference model as

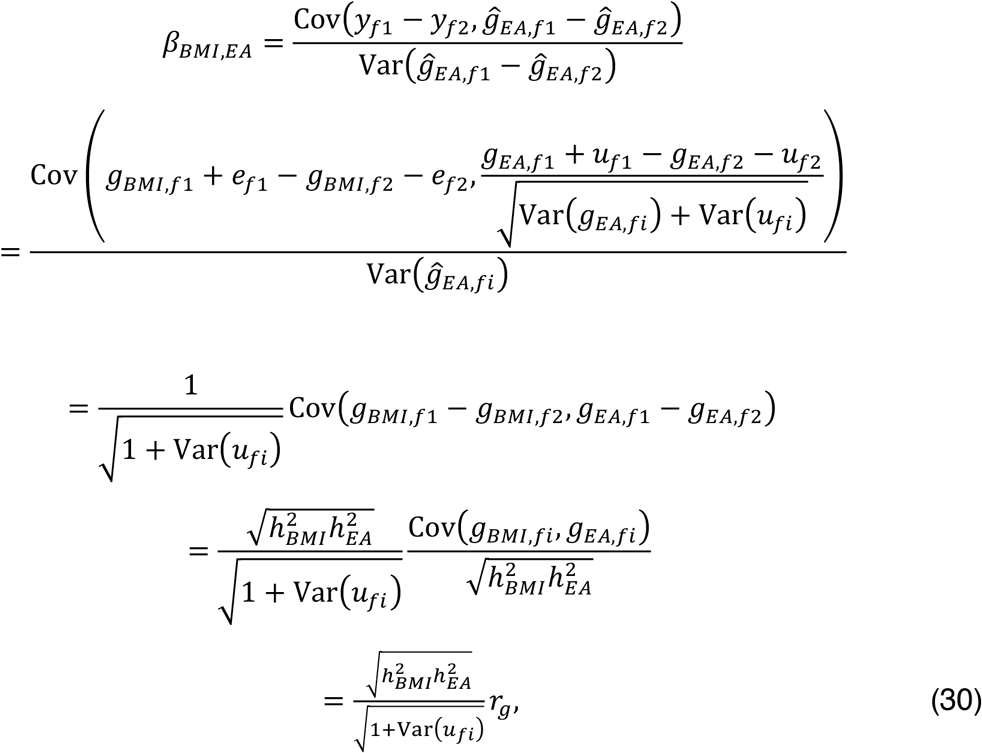

where 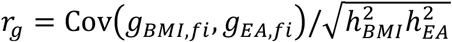 is the genetic correlation. Thus, because 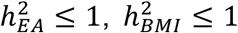, and Var(*u*_*fi*_) > 0, it follows that *β*_*BMI,EA*_ ≤ *r*_*g*_. So the regression coefficient is a lower-bound estimate of the genetic correlation.

### Assortative Mating Simulations

To assess the degree to which GRMA-3^rd^ controls for bias due to assortative mating relative to a population GWAS, we ran a simple simulation with phenotypic sorting on a heritable phenotype. We ran the simulation for several generations until the heritability of the phenotype was in equilibrium. We then estimated (i) the analog of a population GWAS: the association between a single SNP and the heritable phenotype in the full simulated sample; and (ii) the analog of a GRMA-3^rd^ study that only has first cousins in the sample: for each individual in the sample, we selected one of their first cousins at random from the sample and regressed the differences in the cousins’ phenotypes on the difference in the cousins’ genotypes. We estimated the bias as the mean across 10,000 replications of the difference between the estimated association in each analysis and the true, simulated effect size. More details of this simulation are in the **Supplementary Note**.

### Computational Considerations

Since the relatedness groups only need to be formed once and the relatedness-group algorithm is very efficient, most of the computational resources needed are consumed during the estimation process, when each individual genotype entry must be demeaned relative to a relatedness-group mean. Formally, for each individual *i* = 1,2, …, *N* and each SNP *k* = 1,2, …, *K*, the transformation is:

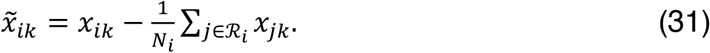

With block processing and parallelization, the step is tractable even for analyses at the scale of the UKB. In our analyses, we found that block sizes of approximately 10,000 SNPs appear to balance memory use and speed close to optimally, yielding a throughput of roughly 0.05 seconds per SNP per core. Processing ∼10 million SNPs therefore requires about 140 CPU-hours. Because computation parallelizes nearly linearly across SNP blocks, a 10M-SNP GRMA analysis can be completed within a few hours in computing environments that support distribution across multiple cores.

## Notes

### Author Declarations

This secondary analysis used de-identified, controlled-access UK Biobank and All of Us data under those programs approved data-use procedures; under University of Southern California policy, the investigators determined that the present study did not constitute human subjects research and therefore did not require USC IRB review or approval.

