## Supplementary Note for "Genomic-Relatedness Matching Expands Population Coverage, Improves Power, and Reduces Bias in Genetic Association Analyses"

**GRMA Study Standard Errors.**

Consider a dataset with $N$ individuals. Imagine we are interested in the effect of some SNP, $k$. Let $\boldsymbol{X}$ denote the $N \times 1$ vector of genotypes of SNP $k$ for each individual, $\boldsymbol{F}$ denote the $N \times 1$ vector of midpoint of each individual's parents' genotypes for SNP $k$, and $\boldsymbol{Y}$ denote the $N \times 1$ vector of each individual's phenotype. (We omit the $k$ subscript here for notational simplicity.) The parameter of interest is the coefficient $\beta$ that would be obtained from the trio-based model:

$$\boldsymbol{Y} = \boldsymbol{X}\beta + \boldsymbol{F}\gamma + \boldsymbol{\epsilon} = \boldsymbol{X}\beta + \boldsymbol{U} \tag{1}$$

where $\boldsymbol{U} = \boldsymbol{F}\gamma + \boldsymbol{\epsilon}$ is the composite residual of unobserved variables. Let $\boldsymbol{\Omega} = \mathrm{Var}(\boldsymbol{U})$ denote the variance-covariance matrix of the residual. Because our data necessarily contain individuals who are related, those individuals may have correlated residuals both due to their genomic relatedness or some degree of a common environment.

The GRMA estimator is based on residualizing the model in Equation (1), within each person's relatedness group (see **Online Methods**). Let $\boldsymbol{R}$ denote the $N \times N$ matrix that residualizes the variables in our samples when left-multiplied by the data. Let $\widetilde{\boldsymbol{Y}} = \boldsymbol{RY}$, $\widetilde{\boldsymbol{X}} = \boldsymbol{RX}$, and $\widetilde{\boldsymbol{U}} = \boldsymbol{RU}$ denote the residualized vectors of $\boldsymbol{Y}$, $\boldsymbol{X}$, and $\boldsymbol{U}$, respectively. This gives us a residualized model

$$\widetilde{\boldsymbol{Y}} = \widetilde{\boldsymbol{X}}\beta + \widetilde{\boldsymbol{U}} \tag{2}$$

which is the model underlying the GRMA estimator, which regresses $\widetilde{\boldsymbol{Y}}$ onto $\widetilde{\boldsymbol{X}}$ using ordinary least squares (OLS). Notice, however, that because the elements of $\boldsymbol{U}$ are not uncorrelated, neither are the elements of $\widetilde{\boldsymbol{U}}$, and therefore conventional OLS standard errors are not appropriate.

We next derive an expression for the sampling variance of a GRMA estimate. Let $\hat{\beta}$ denote the GRMA estimate from an OLS regression as in Equation (2). The variance of this estimate is

$$\begin{aligned}
\mathrm{Var}(\hat{\beta}|\boldsymbol{X},\boldsymbol{R}) &= \mathrm{Var}\left[\left(\widetilde{\boldsymbol{X}}'\widetilde{\boldsymbol{X}}\right)^{-1}\widetilde{\boldsymbol{X}}'\widetilde{\boldsymbol{Y}}\middle|\boldsymbol{X},\boldsymbol{R}\right] \\
&= \mathrm{Var}\left[\left(\widetilde{\boldsymbol{X}}'\widetilde{\boldsymbol{X}}\right)^{-1}\widetilde{\boldsymbol{X}}'\left(\widetilde{\boldsymbol{X}}\beta + \boldsymbol{RU}\right)\middle|\boldsymbol{X},\boldsymbol{R}\right] \\
&= \mathrm{Var}\left[\left(\widetilde{\boldsymbol{X}}'\widetilde{\boldsymbol{X}}\right)^{-1}\widetilde{\boldsymbol{X}}'\boldsymbol{RU}\middle|\boldsymbol{X},\boldsymbol{R}\right] \\
&= \left(\widetilde{\boldsymbol{X}}'\widetilde{\boldsymbol{X}}\right)^{-1}\widetilde{\boldsymbol{X}}'\boldsymbol{R}\mathrm{Var}[\boldsymbol{U}|\boldsymbol{X},\boldsymbol{R}]\boldsymbol{R}'\widetilde{\boldsymbol{X}}\left(\widetilde{\boldsymbol{X}}'\widetilde{\boldsymbol{X}}\right)^{-1} \\
&= \left(\widetilde{\boldsymbol{X}}'\widetilde{\boldsymbol{X}}\right)^{-1}\widetilde{\boldsymbol{X}}'\boldsymbol{R\Omega R}'\widetilde{\boldsymbol{X}}\left(\widetilde{\boldsymbol{X}}'\widetilde{\boldsymbol{X}}\right)^{-1}.
\end{aligned} \tag{3}$$

While $\widetilde{\boldsymbol{X}}$ and $\boldsymbol{R}$ are observed, $\boldsymbol{\Omega}$ is unknown. We therefore estimate this matrix, which we denote $\widehat{\boldsymbol{\Omega}}$, and plug this estimate into Equation (3), similar to the heteroskedasticity-robust, clustered standard error approach[1].

Assuming that $\beta$ is small, it follows that $\boldsymbol{\Omega} = \mathrm{Var}(\boldsymbol{U}) \approx \mathrm{Var}(\boldsymbol{Y})$. This assumption allows us to use the same matrix $\widehat{\boldsymbol{\Omega}}$ for each SNP, which leads to large computational savings. Next, we make two assumptions to estimate $\widehat{\boldsymbol{\Omega}}$. First, we assume that pairs of individuals who are more distant than 3rd-degree relatives have uncorrelated phenotypes. This assumption makes $\widehat{\boldsymbol{\Omega}}$ sufficiently sparse to estimate the standard errors. Second, we assume that the covariance between a pair of individuals depends only on the degree of relatedness between the individuals. For example, this implies that $\boldsymbol{\Omega}_{ij} = \boldsymbol{\Omega}_{k\ell}$ if individuals $i$ and $j$ are 2nd-degree relatives and also individuals $k$ and $\ell$ are 2nd-degree relatives. This also implies that $\boldsymbol{\Omega}$ has identical diagonal entries (i.e., the residuals are homoscedastic) since each person has the same relatedness with themselves.

Under these assumptions, we can estimate the covariance for each degree of relatedness

$$\widehat{\omega}_r = \widehat{\mathrm{Cov}}\left(Y_i, Y_j \;\middle|\; i \text{ and } j \text{ have relatedness } r\right), \tag{4}$$

where $\widehat{\mathrm{Cov}}(\cdot)$ is the sample covariance. Then using these estimates, we define the elements of $\widehat{\boldsymbol{\Omega}}$ to be

$$\widehat{\Omega}_{ij} = \begin{cases} \widehat{\omega}_r & \text{if } i \text{ and } j \text{ have related } r, \text{and} \\ 0 & \text{otherwise.} \end{cases} \tag{5}$$

We then use a simple plug-in estimator, using Equation (3) but substituting $\widehat{\boldsymbol{\Omega}}$ in the place of $\boldsymbol{\Omega}$. We then take the square root of this expression to obtain standard errors:

$$SE(\hat{\beta}) = \sqrt{\left(\widetilde{\boldsymbol{X}}'\widetilde{\boldsymbol{X}}\right)^{-1}\widetilde{\boldsymbol{X}}'\boldsymbol{R}\widehat{\boldsymbol{\Omega}}\boldsymbol{R}'\widetilde{\boldsymbol{X}}\left(\widetilde{\boldsymbol{X}}'\widetilde{\boldsymbol{X}}\right)^{-1}} \tag{6}$$

**Null Simulation to Assess Standard Errors**

Here we describe the simulation used to test whether GRMA's standard errors have correct coverage. The data for this simulation is based on an extended pedigree that contains full-siblings, parent/offspring, 2nd-degree relatives, and 3rd degree relatives in roughly the same proportion that they are observed in the UK Biobank. The pedigree is found in **Supplementary Figure 2**. In this pedigree, we assume that only those with solid dots are observed in our data. We label each solid dot with the degree of relatedness between that person and their closest observed relative: 4 people's closest relative is a full sibling, 1 person's closest relative is a Parent/Offspring, 2 people's closest relative is a 2nd degree, and 7 people's closest relative is a 3rd degree. We assume that 10,000 identical pedigrees exist in our data, which leads to sample sizes approximately equal to those of each relatedness threshold in the UK Biobank data. (See **Table 1**.)

Next, we generate genotypes for 200 SNPs in linkage equilibrium for all individuals in the pedigrees. For individuals with no parents represented in the pedigree, their genotypes are drawn from a Binomial(2,0.4), as they would be in a homogeneous population with no assortative mating and an allele frequency of 0.4 for each SNP. The genotypes for individuals who parents are included in the pedigree are simulated according to the laws of Mendelian inheritance and perfect recombination between each SNP, randomly selecting one allele from each parent at each SNP. Then, genotypes from "unobserved" individuals are dropped from the simulation, leaving 140,000 individuals each with at least one 3rd-degree or closer relative.

Next, we simulate a vector of effect sizes for each SNP. We always fix the effect of the first SNP to be zero, but the effect sizes from the remaining 199 are drawn from a standard normal distribution. The nongenetic component of the phenotype is also drawn from a mean-zero normal distribution, but with variance set such that the heritability of the phenotype is 0.4.

Finally, the genotype of the null SNP, the phenotype, and the known relatedness of each pair of individuals is passed into GRMA to estimate the effect of the SNPs. The estimated effect, standard error, and p-value are all stored in memory.

This simulation is repeated using the same genotype data but newly generated effect sizes and residuals 10,000 times. **Supplementary Figure 1** contains quantile-quantile (QQ) plots of the p-values for the full-sibling, parent-offspring, degree 2, and degree 3 relatedness thresholds.

**The Bias-Variance Tradeoff**

As the relatedness threshold used is relaxed, GRMA faces a bias variance trade-off. (To be more precise, here we define bias as the expected deviation of an estimate from the GRMA estimand using a sibling threshold.) On the one hand, a more permissive relatedness threshold improves precision by increasing the number of individuals included and by increasing the genotypic variance within each relatedness group. On the other hand, greater precision could come at the cost of greater bias. This degree of this trade-off will depend on the phenotype and the sample. For example, if a researcher is studying a phenotype with little confounding, then very permissive thresholds could be used without increasing the bias by much. Alternatively, if there are large parental genetic effects for a particular phenotype in a sample, then the bias from more permissive thresholds may be too large for a particular application.

We have developed a statistic that quantifies the bias-variance tradeoff for a set of $K$ tested SNPs. Letting $\hat{\beta}_k$ denote the GRMA estimate for SNP $k$ and $\beta_k$ denote the sibling-based estimand for that SNP, a researcher would select a threshold that minimizes

$$T(\theta) = \theta\ \sigma_V^2 + (1-\theta)\ \sigma_b^2, \tag{7}$$

where $\sigma_V^2 = K^{-1} \sum_{k=1}^{K} \mathrm{Var}(\hat{\beta}_k)$ is the genome-wide-mean sampling variance, $\sigma_b^2 = K^{-1} \sum_{k=1}^{K} [E(\hat{\beta}_k - \beta_k)]^2$ is the genome-wide-mean squared bias and $\theta \in [0,1]$ is a weight parameter that describes a researchers' aversion to sampling variance relative to bias. A value of $\theta = 1$ would minimize the variance without concern for the bias that might get introduced, and a value of $\theta = 0$ would minimize the bias. A value of $\theta = 0.5$ would correspond to minimizing the mean squared error of the GRMA estimates. The appropriate value of $\theta$ depends on the application.

As described in the **Main Text**, an estimate of $\sigma_V^2$ can be obtained by substituting each summand with its sample analog. However, because the true population parameter $\beta_k$ is unknown, there is not a straightforward way to estimate the bias term, $\hat{\sigma}_b^2$.

First, we define a few terms. By the properties of OLS, the sibling-based GRMA estimate $\hat{\beta}_{\mathrm{sib},k}$ can be expressed as

$$\hat{\beta}_{\mathrm{sib},k} = \beta_k + e_{\mathrm{sib},k}, \tag{8}$$

where $e_{\mathrm{sib},k}$ is the mean-zero estimation error of the sibling-based GRMA estimate, with $\sigma_{\mathrm{sib},k}^2 = \mathrm{Var}(e_{\mathrm{sib},k})$. The potentially biased GRMA estimate would be

$$\hat{\beta}_k = \beta_k + b_k + e_k,$$

where $e_k$ is the mean-zero estimation error of the GRMA estimate, with $\sigma_k^2 = \mathrm{Var}(e_k)$, and $b_k = E(\hat{\beta}_k - \beta_k)$ is the bias of the GRMA estimate relative to the sibling-based estimand. Because $e_{\mathrm{sib}}$ and $e_{\mathrm{GRMA}}$ are just a function of the sample drawn, $\mathrm{Cov}(e_{\mathrm{sib}}, b) = \mathrm{Cov}(e_{\mathrm{GRMA}}, b) = 0$.

Next, consider the expected mean squared deviation of the sibling-based results from the GRMA results:

$$\begin{aligned} \mathrm{E}\left[(\hat{\beta}_k - \hat{\beta}_{\mathrm{sib},k})^2 \mid \beta_k, b_k\right] &= \mathrm{E}\left[(\beta_k + b_k + e_k - \beta_k - e_{\mathrm{sib},k})^2 \mid \beta_k, b_k\right] \\ &= \mathrm{E}\left[(b_k + e_k - e_{\mathrm{sib},k})^2 \mid \beta_k, b_k\right] \\ &= b_k^2 + \mathrm{E}(e_k^2) + \mathrm{E}(e_{\mathrm{sib},k}^2) - 2\,\mathrm{E}(e_k e_{\mathrm{sib},k} \mid \beta_k, b_k) \\ &= b_k^2 + \sigma_k^2 + \sigma_{\mathrm{sib},k}^2 - 2\,\mathrm{E}(e_k e_{\mathrm{sib},k}). \end{aligned} \tag{9}$$

While we have estimates of $\sigma_k^2$ and $\sigma_{\mathrm{sib},k}^2$—namely the squared standard errors if $\hat{\beta}_k$ and $\hat{\beta}_{\mathrm{sib},k}$, respectively—and an estimate of $\mathrm{E}\left[(\hat{\beta}_k - \hat{\beta}_{\mathrm{sib},k})^2 \mid \beta_k, b_k\right]$—the observed value of $(\hat{\beta}_k - \hat{\beta}_{\mathrm{sib},k})^2$—we do not have an estimate of $\mathrm{E}(e_k e_{\mathrm{sib},k})$. Therefore, we next derive an estimator for this term. Recall that the (residulaized) model underlying the GRMA estimator is

$$\tilde{Y}_i = \tilde{X}_{ik}\beta_k + \tilde{U}_{ik} \tag{10}$$

where $\tilde{Y}_i$ denotes the residualized phenotype, $\tilde{X}_{ik}$ denotes the residualized genotype of SNP $k$, and $\tilde{U}_{ik}$ denotes the residualized error term. Let $S$ denote the set of individuals in the sibling sample and let $G$ denote the the set of individuals in the larger GRMA sample. Note that $S \subseteq G$. Importantly, the residualized variable values for the sibling sample are the same as when those siblings are used in the GRMA sample because they are only residualized using the siblings, even if there are more distant relatives in the sample (see **Online Methods**).

Under this notation

$$e_k = \frac{\sum_{i \in G} \tilde{X}_{ik} \tilde{U}_{ik}}{\sum_{i \in G} \tilde{X}_{ik}^2}, \; e_{\mathrm{sib},k} = \frac{\sum_{i \in S} \tilde{X}_{ik} \tilde{U}_{ik}}{\sum_{i \in S} \tilde{X}_{ik}^2}. \tag{11}$$

Therefore

$$\begin{aligned}
\mathrm{E}(e_k e_{\mathrm{sib},k}) &= \mathrm{E}\left[\left(\frac{\sum_{i \in G} \tilde{X}_{ik} \tilde{U}_{ik}}{\sum_{i \in G} \tilde{X}_{ik}^2}\right)\left(\frac{\sum_{i \in S} \tilde{X}_{ik} \tilde{U}_{ik}}{\sum_{i \in S} \tilde{X}_{ik}^2}\right)\right] \\
&= \mathrm{E}\left[\left(\frac{\sum_{i \in S} \tilde{X}_{ik} \tilde{U}_{ik} + \sum_{i \in G-S} \tilde{X}_{ik} \tilde{U}_{ik}}{\sum_{i \in G} \tilde{X}_{ik}^2}\right)\left(\frac{\sum_{i \in S} \tilde{X}_{ik} \tilde{U}_{ik}}{\sum_{i \in S} \tilde{X}_{ik}^2}\right)\right] \\
&= \mathrm{E}\left[\left(\frac{\sum_{i \in S} \tilde{X}_{ik} \tilde{U}_{ik}}{\sum_{i \in G} \tilde{X}_{ik}^2}\right)\left(\frac{\sum_{i \in S} \tilde{X}_{ik} \tilde{U}_{ik}}{\sum_{i \in S} \tilde{X}_{ik}^2}\right)\right] \\
&= \mathrm{E}\left[\left(\frac{\sum_{i \in S} \tilde{X}_{ik} \tilde{U}_{ik}}{\sum_{i \in S} \tilde{X}_{ik}^2}\right)^2 \left(\frac{\sum_{i \in S} \tilde{X}_{ik}^2}{\sum_{i \in G} \tilde{X}_{ik}^2}\right)\right] \\
&= \mathrm{E}(e_{\mathrm{sib},k}^2)\left(\frac{\sum_{i \in S} \tilde{X}_{ik}^2}{\sum_{i \in G} \tilde{X}_{ik}^2}\right) \\
&= \sigma_{\mathrm{sib},k}^2 \left(\frac{\sum_{i \in S} \tilde{X}_{ik}^2}{\sum_{i \in G} \tilde{X}_{ik}^2}\right).
\end{aligned}$$

Substituting this expression into Equation (9) and solving for $b_k^2$ gives us

$$b_k^2 = \mathrm{E}\left[\left(\hat{\beta}_k - \hat{\beta}_{\mathrm{sib},k}\right)^2 \mid \beta_k, b_k\right] - \sigma_k^2 + \sigma_{\mathrm{sib},k}^2 \left[2\left(\frac{\sum_{i \in S} \tilde{X}_{ik}^2}{\sum_{i \in G} \tilde{X}_{ik}^2}\right) - 1\right]. \tag{12}$$

The value $\sigma_b^2$ is just the mean of $b_k^2$ over the set of $K$ SNPs. Substituting the estimates of each the terms of Equation (12) and taking the average over SNPs gives us our estimator

$$\hat{\sigma}_b^2 = \frac{1}{K}\sum_k \left(\left(\hat{\beta}_k - \hat{\beta}_{\mathrm{sib},k}\right)^2 - \hat{\sigma}_k^2 + \hat{\sigma}_{\mathrm{sib},k}^2 \left[2\left(\frac{\sum_{i \in S} \tilde{X}_{ik}^2}{\sum_{i \in G} \tilde{X}_{ik}^2}\right) - 1\right]\right). \tag{13}$$

In the case where we estimate the genome-wide average squared bias of the GWAS estimates, because, by construction, there is no sample overlap between the sibling sample and the GWAS sample, $\mathrm{E}(e_k e_{\mathrm{sib},k}) = 0$, where $e_k$ is the estimation error of the GWAS estimates. As a result,

$$\hat{\sigma}_b^2 = \frac{1}{K}\sum_k \left(\left(\hat{\beta}_k - \hat{\beta}_{\text{sib},k}\right)^2 - \hat{\sigma}_k^2 - \hat{\sigma}_{\text{sib},k}^2\right) \tag{14}$$

As a validation, consider a couple special cases. First, if the GRMA estimate is the sibling estimate. In this case, we would expect $\sigma_b^2 = 0$. Also, $\left(\hat{\beta}_k - \hat{\beta}_{\text{sib},k}\right)^2 = 0$, $\sigma_k^2 = \sigma_{\text{sib},k}^2$, and $\frac{\sum_{i \in S} \tilde{X}_{ik}^2}{\sum_{i \in G} \tilde{X}_{ik}^2} = 1$. Substituting these into Equation (13), we get

$$\hat{\sigma}_b^2 = \frac{1}{K}\sum_k \left(-\hat{\sigma}_{\text{sib},k}^2 + \hat{\sigma}_{\text{sib},k}^2[2(1) - 1]\right) = 0,$$

as expected.

Next, consider the second case where the GRMA sample approaches infinitely large but the sibling sample is finite. Then, $\sigma_k^2 = 0, \mathrm{E}\left[\left(\hat{\beta}_k - \hat{\beta}_{\text{sib},k}\right)^2\right] = b_k^2 + \sigma_{\text{sib,k}}^2$ (because there is no estimation error in $\hat{\beta}_{GRMA}$), and $\frac{\sum_{i \in S} \tilde{X}_{ik}^2}{\sum_{i \in G} \tilde{X}_{ik}^2} = 0$ (because the sum in the bottom approaches infinity). This gives us

$$\begin{aligned} \mathrm{E}(\hat{\sigma}_b^2) &= \mathrm{E}\left[\frac{1}{K}\sum_k \left(\left(\hat{\beta}_k - \hat{\beta}_{\text{sib},k}\right)^2 + \hat{\sigma}_{\text{sib},k}^2[2(0) - 1]\right)\right] \\ &= \frac{1}{K}\sum_k \left(b_k^2 + \sigma_{\text{sib,k}}^2 - \hat{\sigma}_{\text{sib},k}^2\right) = \sigma_b^2, \end{aligned}$$

again as expected.

**Block Jackknife Estimator**

Standard errors on the parameters estimated for the bias-variance trade-off ($\hat{\sigma}_V^2$, $\hat{\sigma}_b^2$, and $\hat{T}(\theta)$) and estimated using a block jackknife procedure over SNPs.

The GRMA summary statistics used in this analysis consist of $M$ SNPs with non-missing estimates in both the sibling-based and GRMA analyses. In a standard jackknife procedure, $M$ estimates, each removing exactly one SNP, and these leave-one-out estimates would be combined to produce standard errors. However, this approach requires that the estimates for each SNP to be uncorrelated. This is not the case due to linkage disequilibrium. So instead, following the approach of LD score regression[2], we employ a block jackknife that groups together SNPs that are potentially correlated.

Specifically, SNPs are ordered by chromosome and physical position and partitioned into $K$ approximately equal-sized, contiguous blocks. In our implementation, $K = 200$. Let $\hat{\sigma}_{b,k}^2$, $\hat{\sigma}_{V,k}^2$, and $\hat{T}_k(\theta)$ denote the corresponding estimates computed using all SNPs except those in block $k$.

The jackknife variance of each parameter is obtained using the standard block jackknife formula. For a generic statistic $\hat{\psi} \in \{\hat{\sigma}_V^2, \hat{\sigma}_b^2, \hat{T}(\theta)\}$, the estimated variance is

$$\widehat{\mathrm{Var}}(\hat{\psi}) \;=\; \frac{K-1}{K}\sum_{k=1}^{K}\left(\hat{\psi}_k - \overline{\psi}\right)^2, \tag{15}$$

where $\overline{\psi} = K^{-1}\sum_{k=1}^{K}\hat{\psi}_{\mathrm{k}}$. Standard errors are reported as the square root of this variance estimate.

**The UK Biobank Data**

*Quality Control of Genetic Data.* We use genotype data from the UK Biobank (UKB), generated and imputed centrally by the UKB following established protocols. Details of genotyping, imputation, and the UKB quality control (QC) pipeline are described in Bycroft et al. (2018)[3]. Briefly, individuals were genotyped in multiple batches using closely related arrays, and extensive marker- and sample-level QC checks were conducted to ensure consistency across batches prior to imputation.

Our analyses begin from the UKB imputed genotype data. At the marker level, we restrict attention to autosomal SNPs that pass standard quality filters. Specifically, we require that SNPs have a minor allele frequency (MAF) greater than 0.01, have a missingness of less than 5%, and an imputation INFO score greater than 0.99, as reported in the UKB imputation quality files. We further require that allele definitions are consistent across data sources and remove SNPs with ambiguous or mismatched allele pairs. After these filters, no duplicate SNP identifiers remain.

To construct the final SNP set used in the GRMA analyses, we intersect the filtered UKB imputed SNPs with a commonly used set of high-quality variants consisting of HapMap3 SNPs and the variants retained by the prediction software SBayesR[4]. This restriction yields a dense set of well-imputed, common SNPs with reliable linkage disequilibrium structure and minimizes sensitivity to imputation artifacts.

We additionally exclude SNPs that fail Hardy–Weinberg equilibrium (HWE) tests in the Unmatched “White British” sample ($p < 10^{-6}$). The reason we restrict to the “white British” sample for this filter is that some SNPs may fail the HWE test because our GRMA analyses are based on a diverse sample, whereas this test is meant to identify SNPs that have distributions consistent with genotyping error. By restricting just to the “white British” sample, we avoid dropping SNPs that are well genotyped but which have heterogeneous allele frequencies across the genetic ancestries represented in the sample. The final set of 828,208 SNPs used in the GRMA analyses is identical across thresholds and phenotypes. **Supplementary Table 3** reports how many SNPs are retained after imposing each of these filters.

*Phenotypes.* Before constructing our phenotypes, we also perform a set of subject-level quality control filters. We exclude individuals who withdrew consent, individuals with ambiguous or discordant sex information, and individuals flagged by the UKB as having poor genotyping quality (including excess heterozygosity, high missingness, sex chromosome aneuploidy, or an unusually large number of close relatives).

We analyze three phenotypes measured in the UK Biobank: body mass index (BMI), height, and educational attainment. Body mass index (BMI) is defined as weight in kilograms divided by height in meters squared and is taken directly from the UKB field measuring BMI at the baseline assessment center visit. Height is defined as standing height in centimeters, also measured at the baseline assessment. For both BMI and height, we restrict the sample to individuals with non-missing measurements and who pass the quality control filters described above.

Educational attainment is constructed from participants' self-reported highest level of education completed. We use the same mapping as done in previous analyses of educational attainment in the UK Biobank[5,6]. This is done by mapping the UKB categorical responses of qualifications received into years of schooling using the International Standard Classification of Education (ISCED) framework. Some categories of qualifications (e.g., the broad category of professional degrees), there is higher variability in the number of years of required to obtain that degree. In such cases, we code a person's years of schooling using their self-reported school-leaving age variable. For more details on how this variable was coded, see Okbay et al.[5].

For all phenotypes, we standardize outcomes within sex prior to analysis. Individuals with missing phenotype values after these restrictions are excluded on a phenotype-by-phenotype basis. Final sample sizes for each phenotype, along with summary statistics, are reported in **Table 1** and **Supplementary Tables 1 and 2**.

**GRMA Summary Statistics**

Manhattan plots for each phenotype and GRMA relatedness threshold can be found in **Supplementary Figures 3-5**. We also use a clumping algorithm to obtain a set of lead SNPs for each phenotype and relatedness threshold. These lead SNPs are reported in **Supplementary Tables 4 and 5**. (Lead SNPs are not reported for educational attainment since no genome-wide significant SNPs were found.)

Our clumping algorithm is the same as those used in previous SSGAC studies[7] and is carried out using Plink. First, the SNP with the lowest p-value is chosen as an index SNP. Second, SNPs within a radius of 248kb from the index variant on the same chromosome whose LD is greater than $r^2 = 0.1$ are assigned to that clump. The algorithm proceeds by iterating through SNPs that are significant at the genome-wide significance level (5e-8) and that have not yet been assigned to a clump, from smallest to largest p-value. In order to calculate pairwise LD, we use the EUR subsample of the 1000 Genomes Phase 3 reference panel. Notably, the samples used in each GRMA study analysis include individuals who have non-EUR genetic ancestries so the LD reference panel may not be representative of the estimations, leading to some lead SNPs being omitted that would be included (or vice versa) if a more appropriate reference panel were used. This concern would in expectation apply to all relatedness thresholds equally, however, so comparisons across thresholds should still be informative about gains in statistical power.

**All of Us Data**

*Quality Control of Genetic Data.* We use genotype data from the All of Us Research Program, generated, imputed, and quality controlled centrally by the program following established pipelines. Details of genotyping, imputation, and primary quality control procedures are described in the previous work[8]. In brief, samples were genotyped using high-density arrays, subjected to extensive sample- and variant-level quality control, and imputed to a large external reference panel.

Our analyses begin from the centrally imputed All of Us genotype data. We restrict attention to autosomal SNPs that pass standard quality filters, closely paralleling those applied in the UK Biobank analyses described above. In particular, we require SNPs to have a minor allele frequency greater than 0.01 and high imputation quality, as measured by the imputation INFO score. We further require consistent allele definitions across datasets and remove SNPs with ambiguous or mismatched allele pairs.

To ensure comparability across cohorts and downstream analyses, we intersect the filtered All of Us imputed SNPs with the same high-quality reference SNP set used in the UK Biobank analyses. This restriction yields a common set of well-imputed, common variants with stable linkage disequilibrium properties and minimizes sensitivity to cohort-specific imputation artifacts.

*Phenotypes.* Before constructing phenotypes, we carry out the same sample-level quality control as we used in the UK Biobank. Both BMI and height are taken directly as reported by *All of Us*.

Educational attainment is constructed from participants' responses to the question "What is the highest grade or year of school you completed?" Responses are mapped into years of schooling using the International Standard Classification of Education (ISCED) framework, following the same conventions used for the UK Biobank analyses and prior large-scale GWAS of educational attainment. A table describing the mapping of participants' responses to this question to a phenotypic value for Educational Attainment can be found in **Supplementary Table 13**.

For all phenotypes, we standardize outcomes within sex prior to analysis. Individuals with missing phenotype values after these restrictions are excluded on a phenotype-by-phenotype basis. Final sample sizes and summary statistics for each phenotype are reported in **Supplementary Table 1**.

**Simulation Study to Assess the Effect of Assortative Mating**

Here we describe the simulation used to test how much bias remains in genetic effect estimates based on cousin differences under a model of phenotypic assortment. In each simulation replicate, we generated genotype and phenotype data for 30 generations of individuals under random Mendelian segregation but phenotypic assortment. Each generation contained 10,000 individuals, split evenly into males and females. Individuals in the founding generation were assigned genotypes at 200 SNPs in linkage equilibrium, with each genotype drawn from a $\mathrm{Binomial}(2, 0.5)$ distribution, implying that each SNP has an allele frequency of 0.5. Letting $v_0^2$ denote the assumed genetic variance under random mating, a vector of SNP effect sizes was drawn from a $\mathrm{N}(0, v_0^2/[200 \times 2 \times .5 \times (1 - .5)])$ distribution, such that there is a constant expected contribution of each SNP to the heritability. The exception is that the effect size of the first SNP is always set to exactly $\sqrt{v_0^2/[200 \times 2 \times .5 \times (1 - .5)]}$. This is done because this SNP will be used to assess the bias under different estimation strategies so we would like the baseline effect size to be the same across all replications.

The value of $v_0^2$ is selected to correspond to a target value of equilibrium heritability after many generations of phenotypic assortment, where mating pairs have a constant phenotypic correlation in each generation. It has been shown[9,10] that for a target heritability of $h^2$ and a parental phenotypic correlation of $r$,

$$v_0^2 = h^2(1 - rh^2).$$

Each person's phenotype $y_i$ was generated as the sum of an additive genetic component and an independent environmental residual. Specifically, the genetic component was defined as the standardized genotype matrix multiplied by the vector of SNP effects, and the residual was drawn from a $\mathrm{N}(0, 1 - h^2)$ distribution.

Assortative mating was induced by first randomly assigning individuals within each generation to male and female groups, and creating a sorting variable equal to

$$s_i = \sqrt{r}\, y_i + \sqrt{1 - r}\, e_i$$

where $e_i$ is a mean-zero normal variable with variance equal to the sample variance of $y_i$ in that person's generation. (In equilibrium, $y_i$ has a variance of one, but it is less than one in earlier generations.) Males and females are matched ordinally on $s_i$, which leads to a near perfect parental correlation of $s_i$ and a parental correlation of $r$ for $y_i$. Each mating pair produced two offspring, and offspring genotypes were generated SNP-by-SNP by randomly transmitting one allele from each parent. This procedure was repeated for 30 generations.

At the end of the final generation, we estimated the effect of the focal SNP using three estimators. First, we estimated the ordinary population association by regressing phenotype on the SNP. Second, we estimated a sibling-difference association by regressing the phenotypic difference between siblings onto the genotypic difference. Third, we estimated a cousin-difference association by randomly selecting one cousin for each individual (when at least one cousin was available) and regressing the phenotype difference between the individual and that cousin on the corresponding genotype difference. Thus, the cousin-difference estimator was based only on variation within cousin pairs in the final generation.

For each replicate, we recorded the difference between the estimated coefficient on the focal SNP and its true simulated effect size. We repeated the entire simulation 10,000 times and summarized the mean bias for the population, sibling-difference, and cousin-difference estimators. We also computed the ratio of the mean cousin-difference bias to the mean population-association bias as a measure of how much bias due to assortative mating remains when using a cousin difference design.

The values for $h^2$ and $r$ were taken from the literature for BMI, educational attainment, and height. Sibling-based estimates of heritability have been estimated[11] as 0.58 for BMI, .0.08 for educational attainment, and 0.64 for height. Mate correlations have been estimated[12] to be 0.26 for BMI, 0.47 for educational attainment, and 0.24 for height. We use these values in our simulation.
