## Supplementary Figures for "Genomic-Relatedness Matching Expands Population Coverage, Improves Power, and Reduces Bias in Genetic Association Analyses"

**Supplementary Figure 1.** The plot depicts QQ plots of p-values for a null SNP in 10,000 replications of simulated data. (See Online Methods and Supplementary Note.) The y-axis corresponds to the observed $-log_{10}(p)$, and the x-axis corresponds to the expected $-log_{10}(p)$, under the null distribution. Each panel contains the p-values for a GRMA studies at a different relatedness thresholds. **(a)** Full siblings (GRMA-S), **(b)** Parent-offspring (GRMA-PO), **(c)** $2^{nd}$-degree (GRMA-$2^{nd}$), **(d)** $3^{rd}$-degree (GRMA-$3^{rd}$).

**(a)** GRMA-S

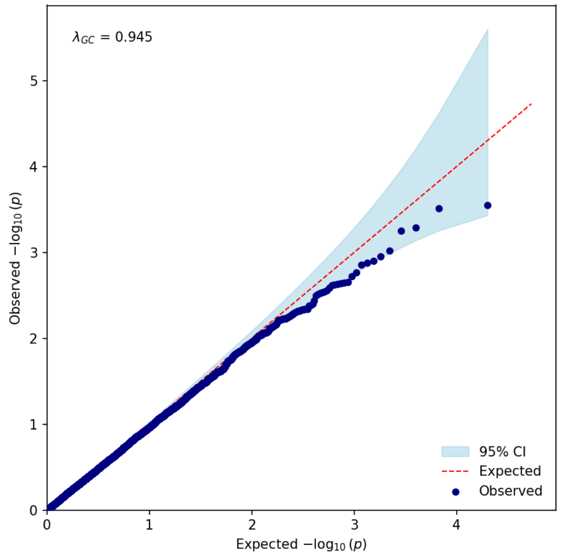


**(b)** GRMA-PO

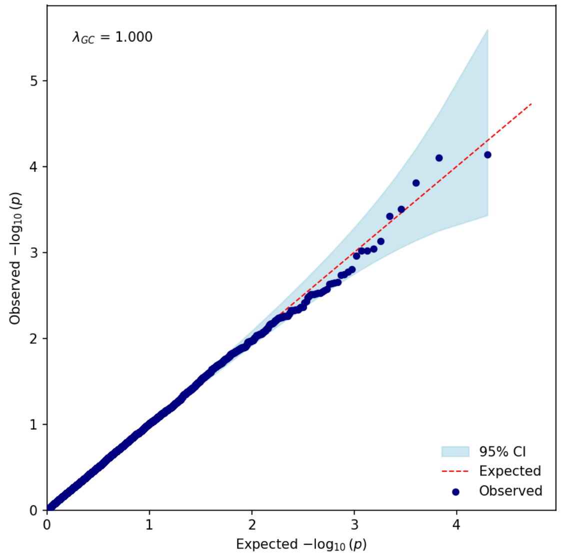


**(c)** GRMA-$2^{nd}$

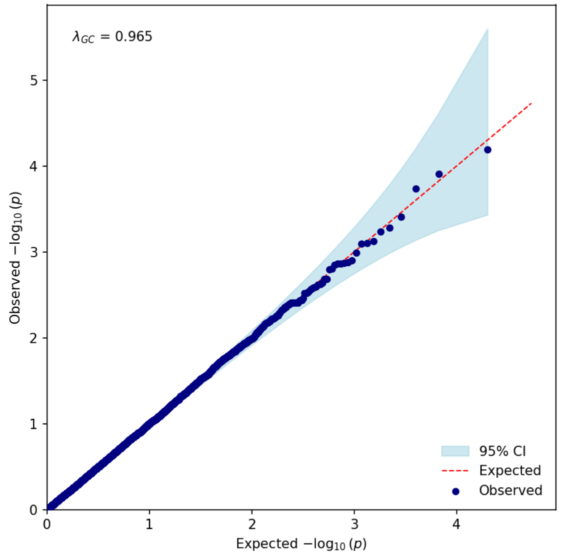


**(d)** GRMA-$3^{rd}$

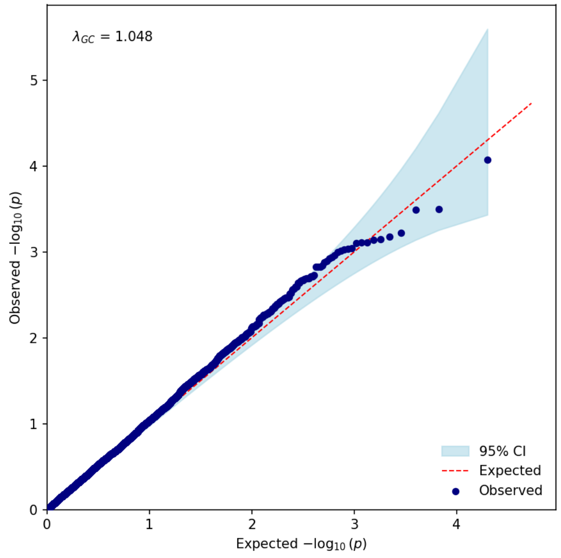

**Supplementary Figure 2.** Pedigree used in the null simulation to assess the coverage of the standard errors. In this simulation, we simulate genotypes for every person in this pedigree according to the laws of Mendelian inheritance, assuming a random mating population. Hollow circles correspond to "unobserved" individuals whose genotypes are deleted before analyzing the simulated data; solid circles correspond to "observed" individuals who are analyzed. Each solid circle is labeled with degree-relatedness of their closest observed relative (FS: full sibling; PO: parent/offspring; 2: second-degree relative; 3: third-degree relative). Within each replication of the simulation, this pedigree was simulated 10,000 times in order to approximate the GRMA sample sizes in the UK Biobank. (See Online Methods and Supplementary Note.)

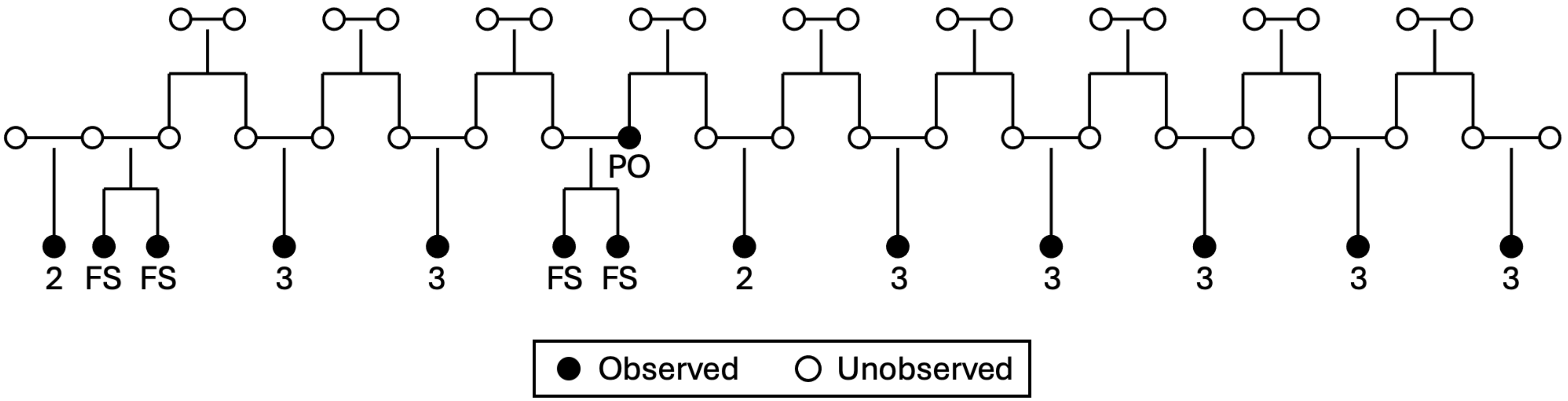

**Supplementary Figure 3**: Manhattan plots for BMI. Each panel corresponds to a different relatedness threshold. The *x* axis is chromosomal position, and the *y* axis is the significance on a –log10 scale. The dashed line marks the threshold for genome-wide significance ($P$ = 5 × $10^{-8}$).

**(a)** GRMA-S

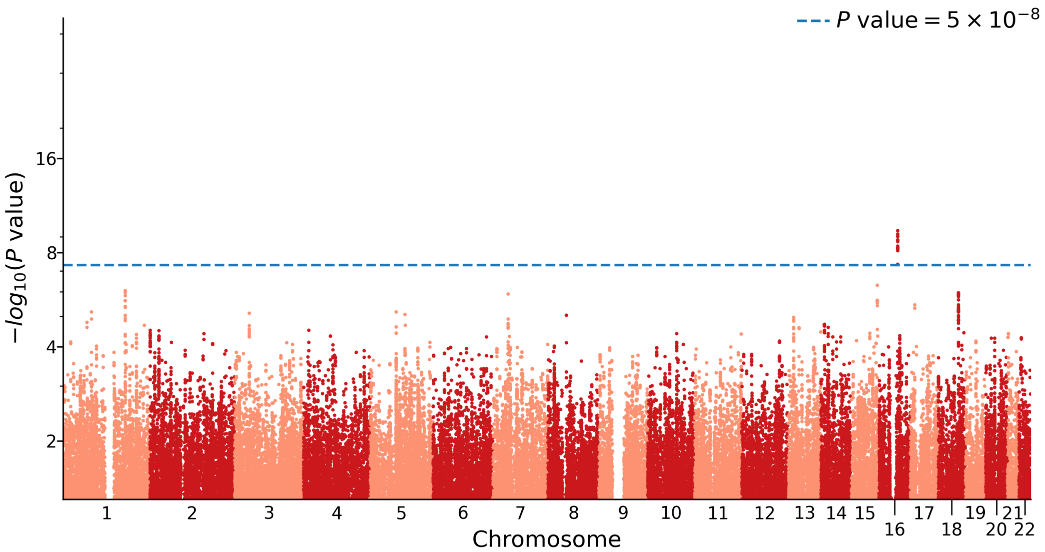


**(b)** GRMA-PO

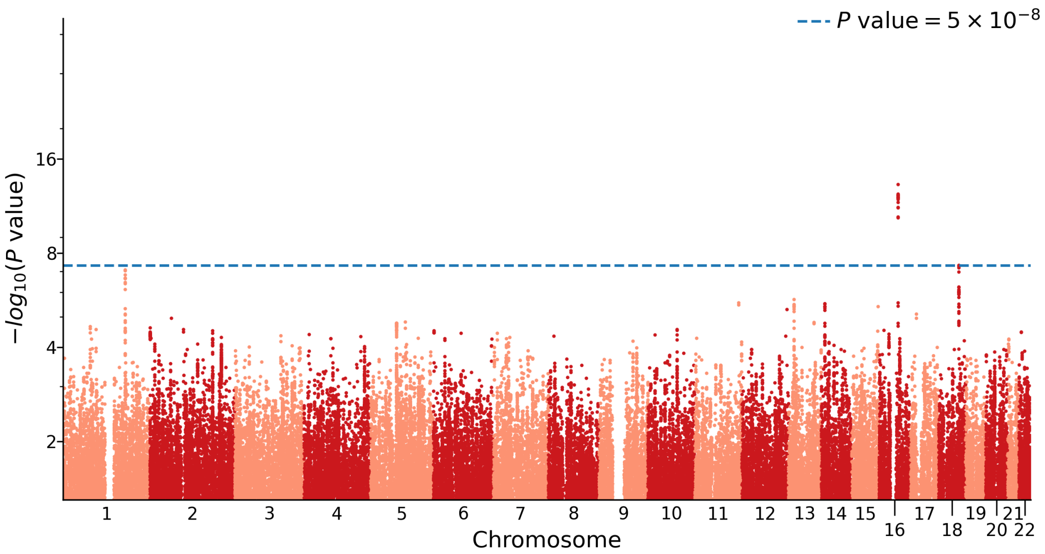

**(c)** GRMA-2[nd]

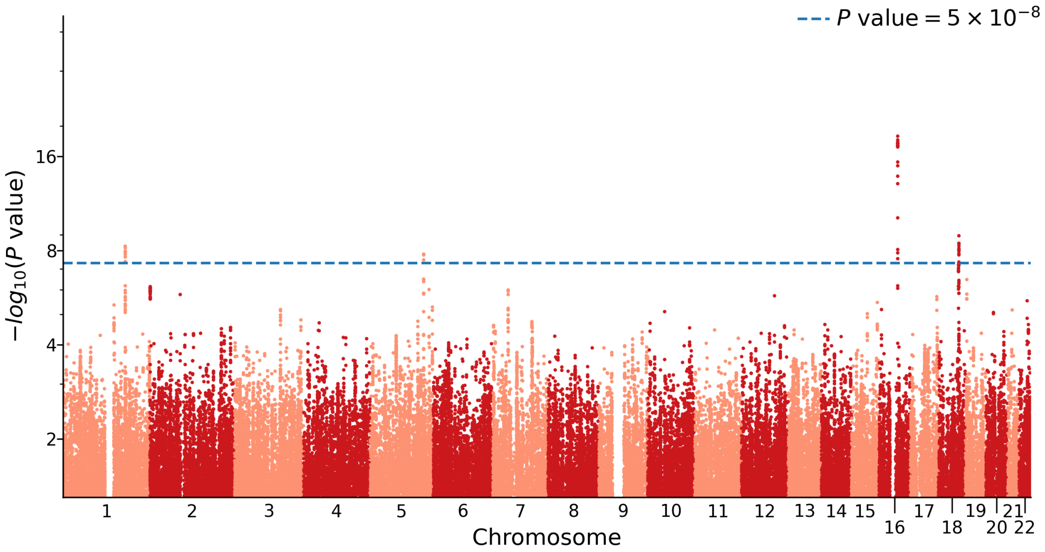


**(d)** GRMA-3[rd]

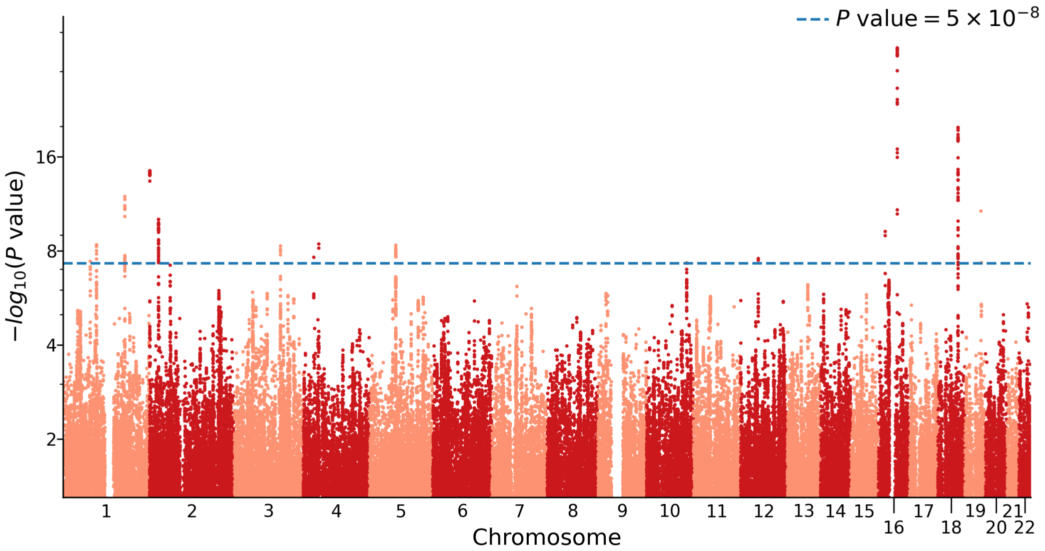

**Supplementary Figure 4**: Manhattan Plots for Height. Each panel corresponds to a different relatedness threshold. The *x* axis is chromosomal position, and the *y* axis is the significance on a –log10 scale. The dashed line marks the threshold for genome-wide significance ($P = 5 \times 10^{-8}$).

**(a)** GRMA-S

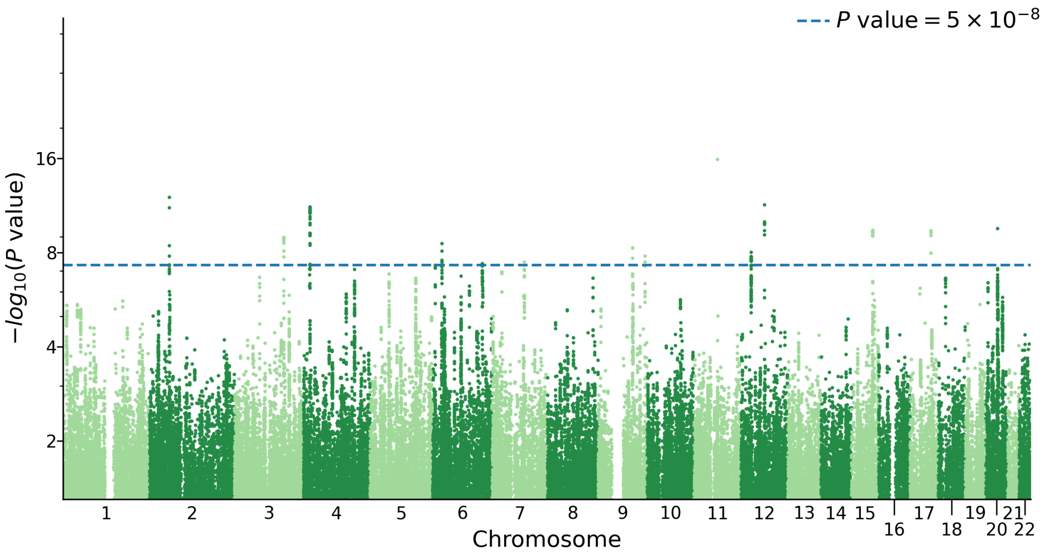


**(b)** GRMA-PO

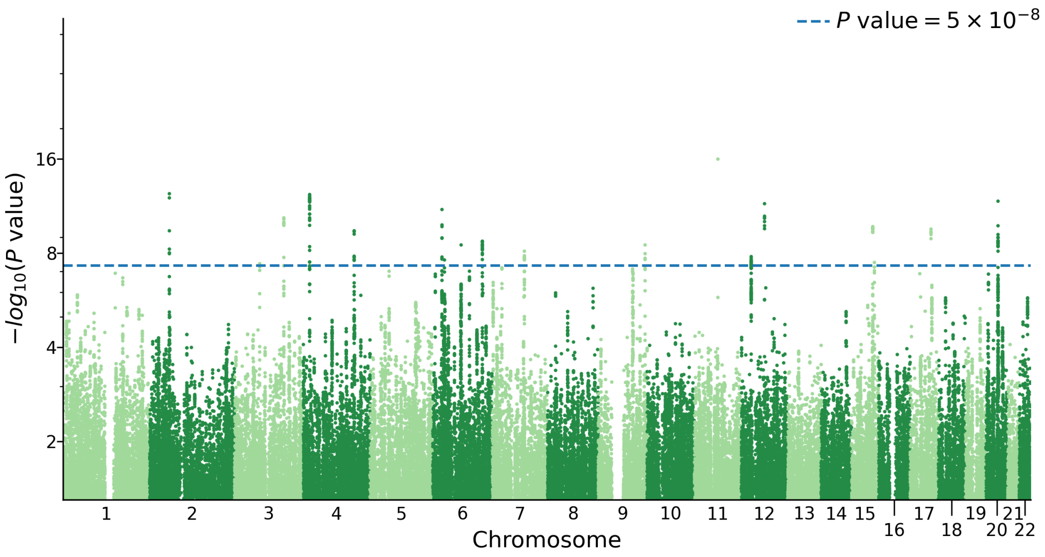

**(c)** GRMA-2nd

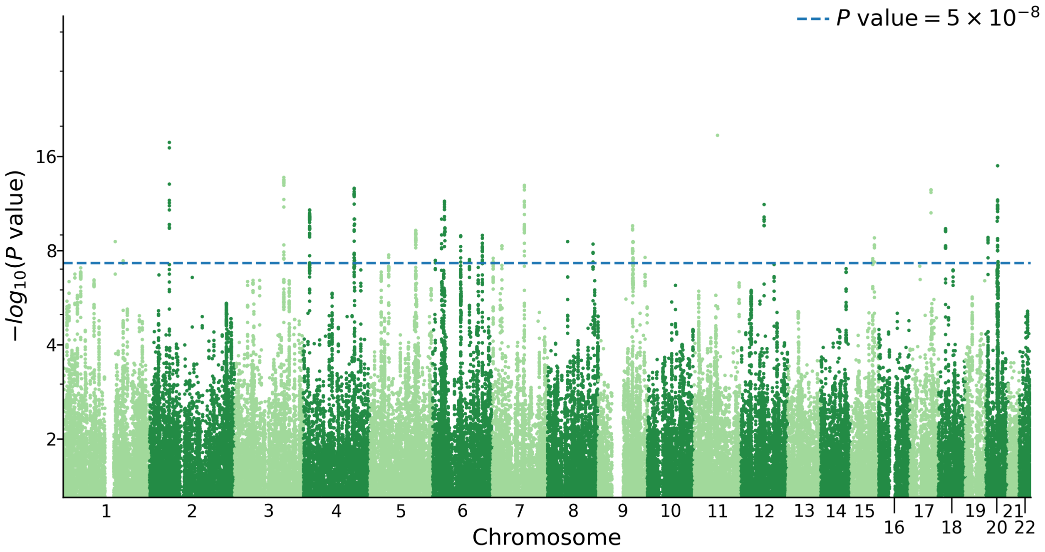


**(d)** GRMA-3rd

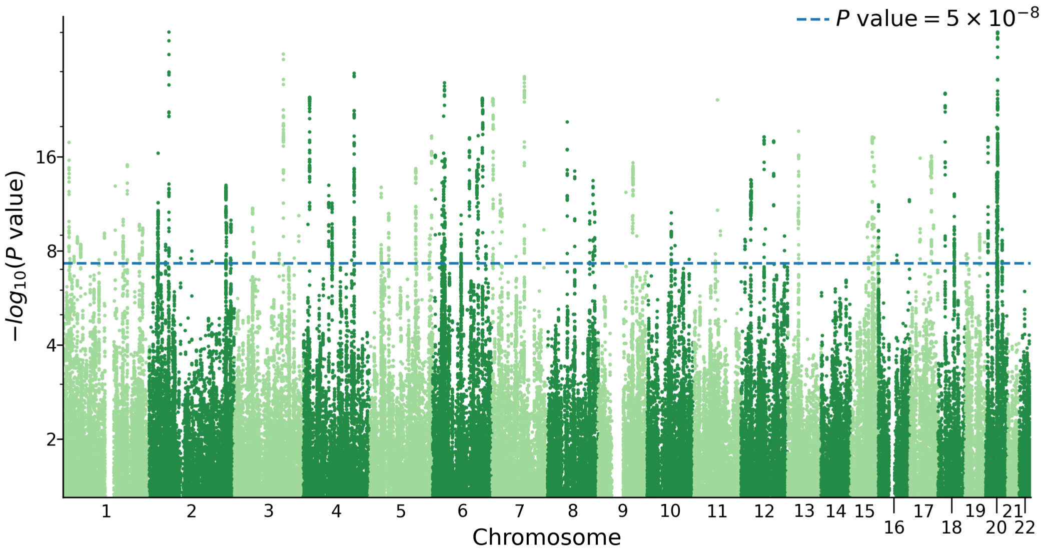

**Supplementary Figure 5**: Manhattan Plots for Educational Attainment. Each panel corresponds to a different relatedness threshold. The *x* axis is chromosomal position, and the *y* axis is the significance on a –log10 scale. The dashed line marks the threshold for genome-wide significance ($P = 5 \times 10^{-8}$).

**(a)** GRMA-S

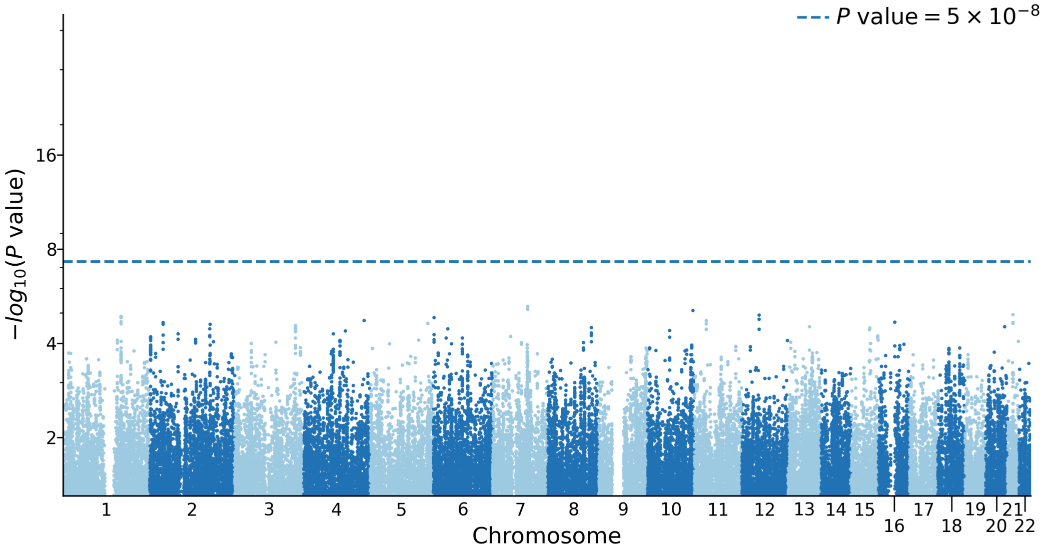


**(b)** GRMA-PO

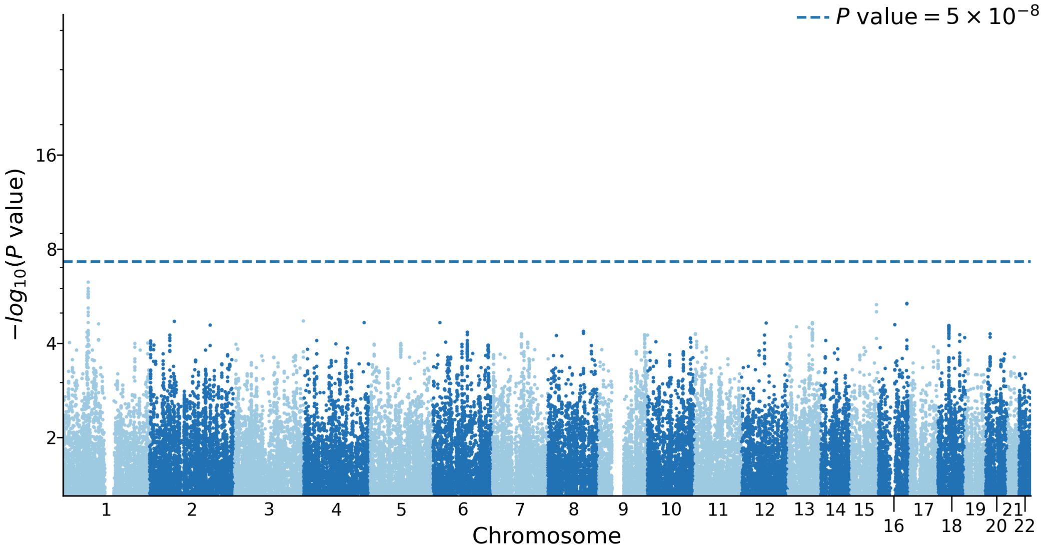

**(c)** GRMA-2$^{nd}$

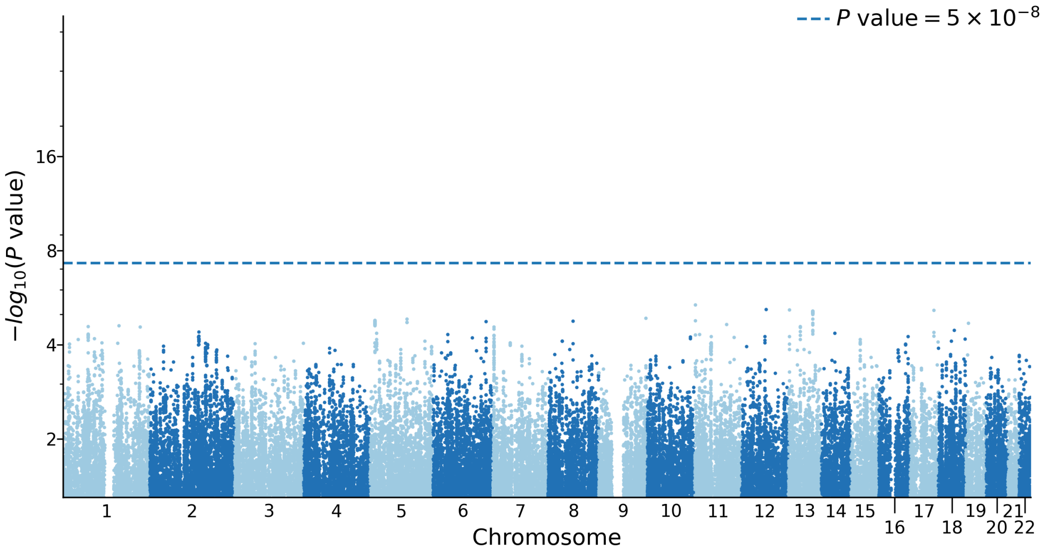


**(d)** GRMA-3$^{rd}$

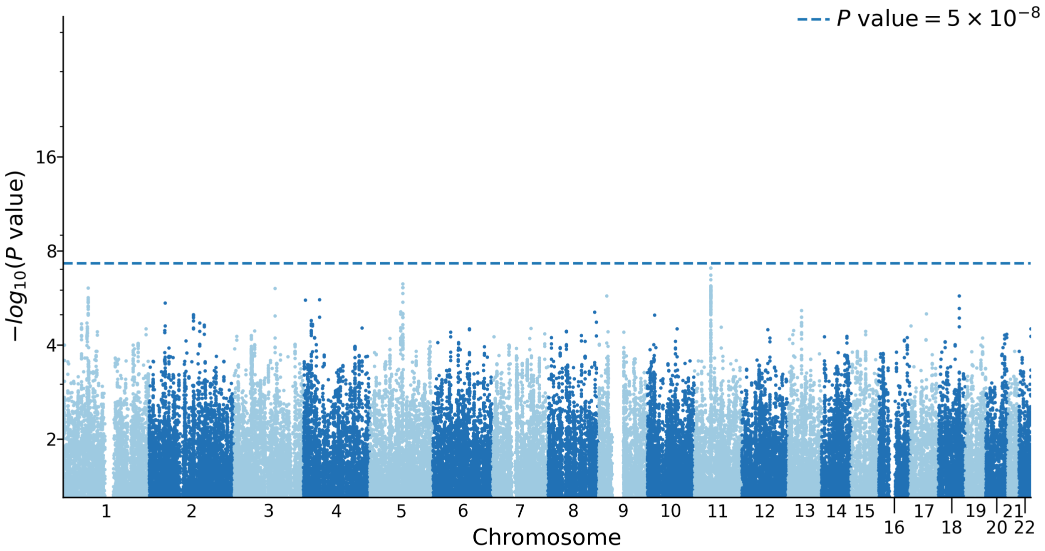
